# Simultaneous detection and quantification of multiple pathogen targets in wastewater

**DOI:** 10.1101/2023.06.23.23291792

**Authors:** Gouthami Rao, Drew Capone, Kevin Zhu, Abigail Knoble, Yarrow Linden, Ryan Clark, Amanda Lai, Juhee Kim, Ching-Hua Huang, Aaron Bivins, Joe Brown

## Abstract

Wastewater-based epidemiology has emerged as a critical tool for public health surveillance, building on decades of environmental surveillance work for pathogens such as poliovirus. Work to date has been limited to monitoring a single pathogen or small numbers of pathogens in targeted studies; however, few studies consider simultaneous quantitative analysis of a wide variety of pathogens, which could greatly increase the utility of wastewater surveillance. We developed a novel quantitative multi-pathogen surveillance approach (35 pathogen targets including bacteria, viruses, protozoa, and helminths) using TaqMan Array Cards (TAC) and applied the method on concentrated wastewater samples collected at four wastewater treatment plants in Atlanta, GA from February to October of 2020. From sewersheds serving approximately 2 million people, we detected a wide range of targets including many we expected to find in wastewater (e.g., enterotoxigenic *E. coli* and *Giardia* in 97% of 29 samples at stable concentrations) as well as unexpected targets including *Strongyloides stercoralis* (a human threadworm rarely observed in the USA). Other notable detections included SARS-CoV-2, but also several pathogen targets that are not commonly included in wastewater surveillance like *Acanthamoeba* spp., *Balantidium coli*, *Entamoeba histolytica*, astrovirus, norovirus, and sapovirus. Our data suggest broad utility in expanding the scope of enteric pathogen surveillance in wastewaters, with potential for application in a variety of settings where pathogen quantification in fecal waste streams can inform public health surveillance and selection of control measures to limit infections.

## Introduction

Wastewater-based epidemiology (WBE) incorporates a range of tools intended to complement traditional public health surveillance, optimally providing timely and actionable data on pathogens circulating in populations of interest. Historically, wastewater monitoring has been used as a surveillance tool for individual pathogens including poliovirus[1,2], hepatitis A[3], *Vibrio cholerae*[4], *Salmonella enterica* serotype Typhi [5] as well as for chemical analytes (e.g., drug use) [6]. This strategy has gained global prominence in the detection and quantification of SARS-CoV-2 RNA in wastewater[7–9], specifically focusing on community prevalence[7,10,11], apparent trends in infections over time and space[12], and emerging variants[13,14]. Advantages and limitations of wastewater as a surveillance matrix have been widely discussed since 2020[15–17].

The need to expand wastewater monitoring to screen multiple pathogens or variants is a valuable approach to better understand the possibility of emerging pathogens or circulating strains in a particular population. In addition to a rapidly expanding array of sequencing techniques to more completely characterize microbial composition of environmental samples, more sensitive quantitative or semi-quantitative multiple-target detection approaches exist [18,19] and some have been subjected to cross-method comparisons for pathogen detection and quantification [20–22]. Such tests could complement the highly sensitive and precisely quantitative emerging digital PCR techniques now considered the gold standard for single-pathogen detection in wastewater, either as a screening method as a precursor to more in-depth work on targets of interest or to gain information on a wide range of pathogens of interest. Emerging and re-emerging infectious diseases[23] – including those with pandemic potential[24] – represent ongoing risks to society, and wastewater surveillance can fill critical gaps in data to inform public health responses[25].

Based on the demonstrated potential for WBE to complement traditional diagnostic public health surveillance for a diverse array of pathogens, we implemented a customized multi-parallel molecular surveillance tool for simultaneous detection and quantification of 35 common pathogenic bacteria, viruses, protozoa, and helminths in wastewater. Such approaches can expand the existing WBE platform by screening for many more pathogens – including rare or emerging microbes of interest – enhancing monitoring to inform public health response. We demonstrate the utility of this method in an analysis of primary untreated influent samples from four wastewater treatment plants in metro Atlanta, Georgia, USA.

## Materials & Methods

### Sample Collection

We collected one-liter primary influent grab samples (n=30) in high-density polyethylene (HDPE) plastic bottles from four wastewater treatment plants (anonymized as WWTP A, B, C, D) in Atlanta, GA between March 20^th^, 2020 - November 5^th^, 2020 between 9:30 AM—11 AM. We obtained permission for sample collection from each WWTP manager prior to sampling. Flow values from the WWTPs ranged from 14 – 80.2 million gallons per day. All samples were transferred to the laboratory on ice and stored at −80°C until further processing was completed. Initial sample processing began on November 8^th^, 2021. Frozen samples were thawed in a 5L bucket of water located in a 4°C walk-in fridge for up to 3 days or until thawed. Samples were then recorded for temperature and pH, and a 50 mL aliquot was taken for total suspended solids measurements (S1 Table). Each 1L sample was spiked with 10 µL of Calf-Guard (Zoetis) resuspended vaccine, containing attenuated bovine coronavirus (BCoV), and 10 µL of MS2 (10^5^/µL), which served as the process recovery controls. A 1:100 ratio of 5% Tween 20 solution was added to the sample bottle as recommended by InnovaPrep for processing wastewater samples [26]. A graduated 1L bottle was used as a reference for the total volume in each sample bottle. Samples were mixed by inverting the bottle 3-4 times. A subset of samples (n=4) were processed using three different methods to establish a reasonable workflow for the remaining samples: (1) direct extraction, (2) InnovaPrep Concentrating Pipette (CP) Select, and (3) skim milk flocculation (SMF).

### Sample Processing

#### Direct Extraction

We directly extracted 200 µL of wastewater influent into the DNeasy PowerSoil Pro Manual extraction kit (Qiagen, Hilden, Germany). Technical representatives indicated kits co-purify DNA and RNA and others have compared DNA kits with DNA + RNA kits with similar performance [27].

#### InnovaPrep Concentrating Pipette

150 mL from the wastewater influent sample was transferred into a 500 mL conical centrifuge tube. Samples were centrifuged for 20 minutes at 4800 x g. The 500 mL conical tube was placed under the CP Select, and the fluidics head lowered into the sample. The sample supernatant was filtered using a 0.05 µm unirradiated hollow-fiber CP tip and eluted using the InnovaPrep FluidPrep Tris elution canister. Processing times and eluted volumes were recorded. For each day samples were run, one negative control consisting of 100 mL of DI water was also filtered and processed.

#### Skim Milk Flocculation

With the remaining wastewater sample, we proceeded to use the SMF method[28]. We combined 1 mL of a 5% skimmed milk solution per 100 mL of wastewater sample (average volume = 750 mL) and adjusted the pH of the skimmed-milk-wastewater solution between 3.0 – 4.0 using 1M HCl. Samples were placed on a shaker plate at room temperature (20-25°C) at 200 RPM for two hours. After shaking, samples were centrifuged at 3500 x g at 4°C for 30 minutes. The supernatant was discarded and the pellet was archived at −80°C until two batch extractions of 15 samples were completed within one week.

A subset of 4 samples were directly extracted and the TaqMan Array Card (TAC) results from CP, SMF, and the direct extractions were compared to determine an optimal concentration method prior to full scale downstream processing. Additional details can be found in S2 Table. In the methods trial, SMF resulted in greater number of pathogen detections and was therefore used for the subsequent full-scale analyses. In the SMF workflow, skim milk pellets were processed for RNA using the Qiagen DNeasy PowerSoil Pro manual extraction kit. One extraction blank was run using nuclease-free water for each batch of sample extractions. Extracts were placed in the −80°C freezer until reverse transcriptase real-time (quantitative) polymerase chain reaction (RT-qPCR) or digital PCR (dPCR) processing followed within one week. Skim milk pellets were run on TAC with 7% in duplicate. All CP eluants were extracted for RNA using Qiagen AllPrep PowerViral manual kits following manufacturer instructions to be further processed using dPCR. CP and dPCR were used for process controls and fecal indicators in the full-scale analyses.

#### Molecular Analysis

Two PCR platforms were used to process extracts, the first was an RT-qPCR QuantStudio (QS) 7 Flex (ThermoFisher Scientific, Waltham, MA) and the second a dPCR QIAcuity Four (Qiagen, Hilden, Germany). All skim milk pellets were analyzed using the QS7 Flex. The QS7 works in conjunction with a custom TAC, which is prespecified with lyophilized primers and probes for 35 enteric pathogen targets (see S3 Table). The card was designed to include bacterial, viral, protozoan, and helminth targets that may be circulating in the United States as well as the leading etiologies of diarrhea among children globally [29,30]. Cq values < 40 were considered positive for the target and confirmed through clear amplification signals in the amplification and multicomponent plots. We prepared our TAC by combining 38 µL of template with 62 µL of AgPath-ID One-Step RT-PCR Reagents (Applied Biosystems) and assessed TAC performance through an 8-fold dilution series (10^9^-10^2^ gene copies/reaction) using 2 plasmids (one for DNA and one for RNA targets) that were linearized, transcribed, cleaned, and quantified as described in [29]. The samples were analyzed in single, not replicates on the same TAC. Additional MIQE details are found in S4 Table. All CP eluant samples were analyzed using the dPCR QIAcuity Four platform (Qiagen, Hilden, Germany). On the dPCR platform previously designed and optimized multiplex assays were used for bovine coronavirus (BCoV), pepper mild mottle virus (PMMoV), and human mitochondrial DNA (mtDNA)[31] (see S5 Table, S1 Text, and S1 Fig)). Gene copy concentration results for PMMoV and mtDNA were used as normalization markers for the TAC pathogen data so that we divided the sample gene copy concentrations/liter by the normalization marker gene copy concentrations/liter.

#### Data Analysis

When multiple gene targets for a single microbial taxon was detected, we used the highest concentration gene target to calculate summary statistics and supported figures. We used R Studio version 4.2.1 and specific R packages to complete all data cleaning (dplyr v1.1.2), analyses (janitor v2.2.0, gtsummary v1.7.1) and generate graphs (ggplot2 v3.4.2). All TAC data was analyzed using QuantStudio Design and Analysis Real-Time PCR software (v2.6.0, Thermo Fisher Scientific). Equivalent sample volumes (ESV) have previously been described as the original sample volume processed and analyzed in a PCR reaction[32]. Here, we calculated ESVs using the following equation:

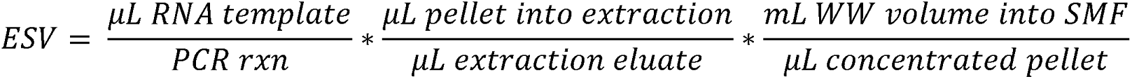

The 95% limit of detections (LODs) were calculated for each assay using probit models[33]. We translated these 95% analytical LODs (aLODs) into a 95% matrix LOD (mLOD) using the following equation and the previously calculated effective volumes for SMF:

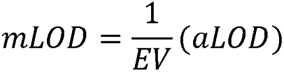

## Results

TAC results were generated using skim milk pellets extracted by the PowerSoil Pro Manual kit to process the influent samples. The average SMF pellet was 2.2 mL and the average wastewater influent processed for SMF was 688 mL. Supplemental data on any other method performed (direct extraction or InnovaPrep CP pellet) is provided in S2 Table and S2-S3 Figs.

### Enteric Pathogen Measurement by Skim Milk Flocculation

The log_10_-transformed gene copy concentrations by pathogen class and specific enteric pathogen (Fig 1) demonstrates the wide range of pathogens detected in Atlanta wastewater influent (n=30). Enteric bacteria, specifically enterotoxigenic *E. coli* (ETEC), were detected most frequently and at higher gene copy concentrations compared to helminths and viruses. Notable protozoan detections were *Acanthamoeba* spp. (28/30), *Balantidium coli* (29/30)*, Entamoeba* spp. (29/30), and *Giardia* spp. (29/30). While virus detections were relatively lower than protozoan detections, astrovirus (26/30), norovirus GI/GII (28/30), and sapovirus (7/30) were detected in the processed samples. Additional comparison of prevalence of pathogens by wastewater treatment plant are detailed in Table 2 with Plant C representing the most samples processed (n=21). S4 Fig demonstrates the log_10_ gene copies per liter of wastewater influent stratified by gene targets. Interestingly, with the CP samples we detected *Strongyloides stercoralis* in one wastewater sample (S2 Fig and S6 Table).

**Fig 1.**
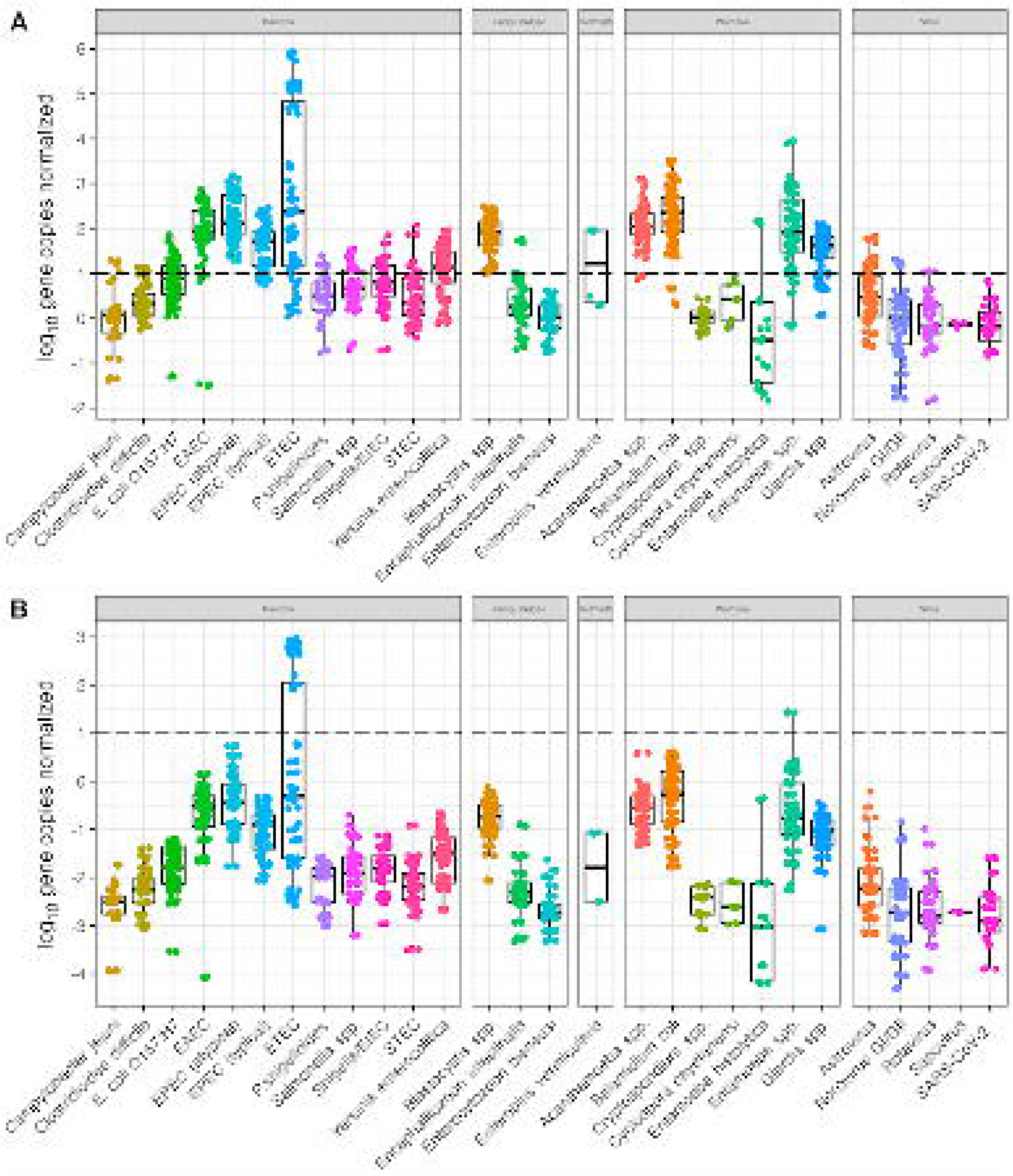
Log_10_ concentrations of enteric pathogens per liter of wastewater influent using the SMF method and PowerSoil Pro Manual extraction.

**Table 2.**
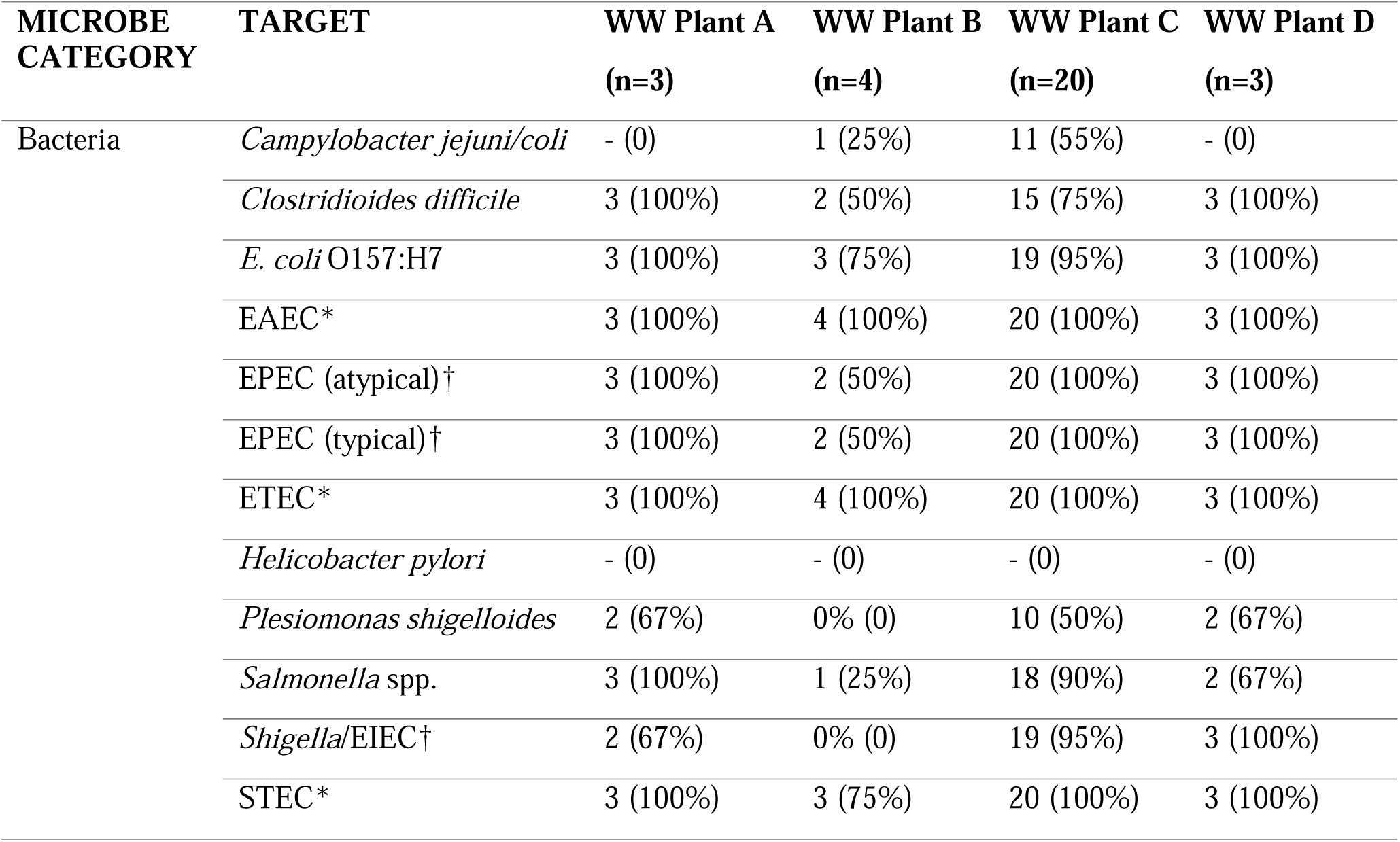
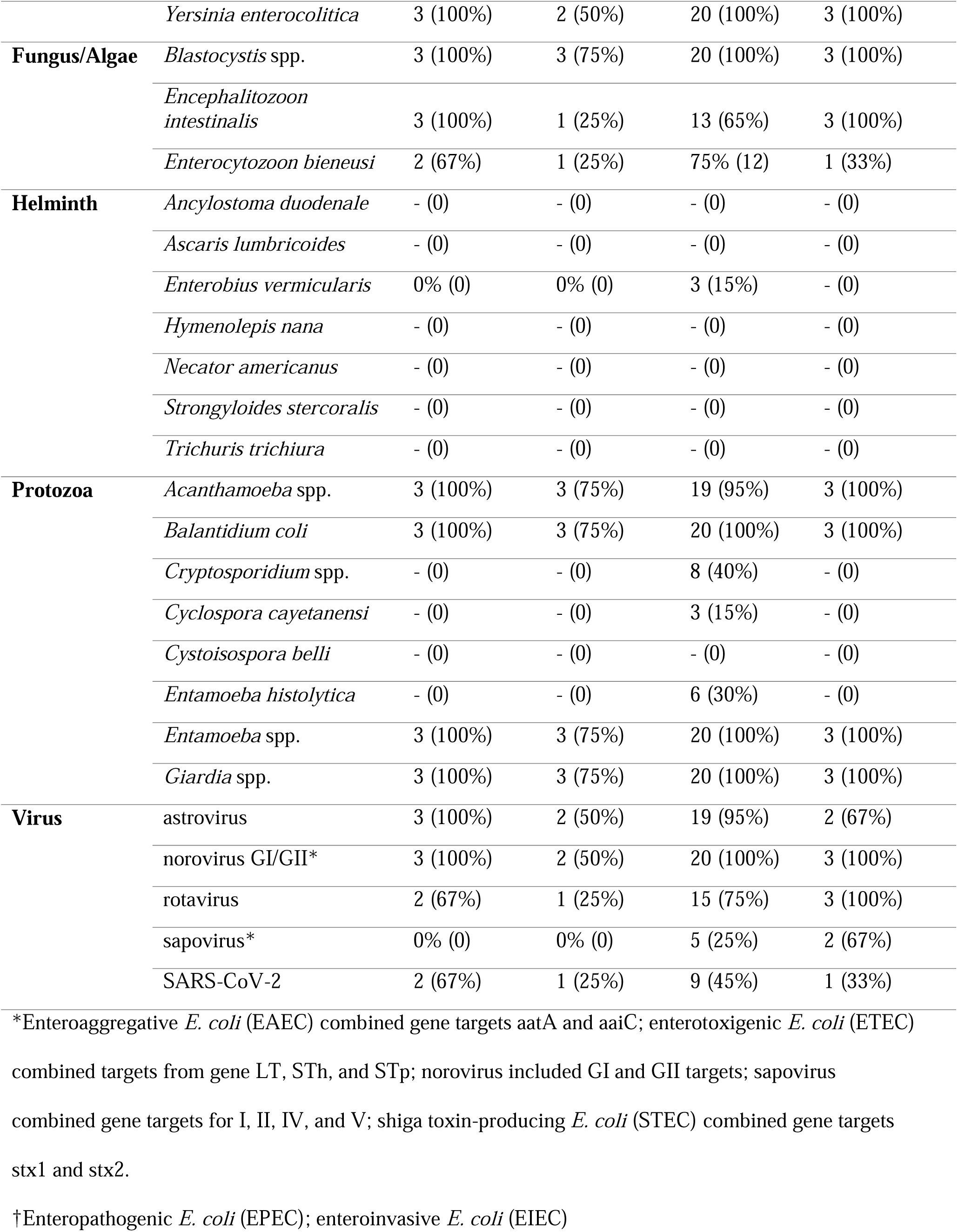
Prevalence of pathogens [n by column (%)] detected in wastewater influent from four treatment plants in Atlanta, Georgia – using SMF method.

Of the SMF samples, the bacterial targets of highest concentration were ETEC and enteropathogenic *E. coli* (EPEC - atypical), whereas viral targets were mainly astrovirus and norovirus GI/GII. Somewhat unexpected protozoan targets detected were *Cyclospora cayetanensi* (3/30) and *Entamoeba histolytica* (6/30). Both *Cryptosporidium* spp. and *Giardia* spp. were detected at means of 5.0 log_10_ and 6.5 log_10_, respectively. Of the total samples, we detected SARS-CoV-2 RNA in 50% of samples (n=15) at concentrations between 3.0 log_10_—6.0 log_10_ gene copies per liter of wastewater influent.

### dPCR for Concentrating Pipette and normalization markers

A total of n=30 CP samples were processed for PMMoV, mtDNA, and BCoV. Fig 2 demonstrates the log_10_ gene copies per liter of wastewater influent and indicates PMMoV concentrations exceed mtDNA concentrations. The average concentrations for BCoV dPCR reactions was 43.3 gene copies (gc)/μL, PMMoV was 1602 gc/μL, and mtDNA was 4.33 gc/μL. The average concentrations of log_10_ gene copies/liter per reaction of wastewater was 5.2 x 10^4^ for mtDNA and 1.9 x 10^7^ for PMMoV. All positive controls and non-template controls performed without suspicion and additional details on control performance is included in S2 Text and in the dMIQE checklist (S7 Table). Additionally, BCoV as a process control yielded a 29% average recovery with a standard deviation of 28, with recovery by sample available as S8 Table.

**Fig 2.**
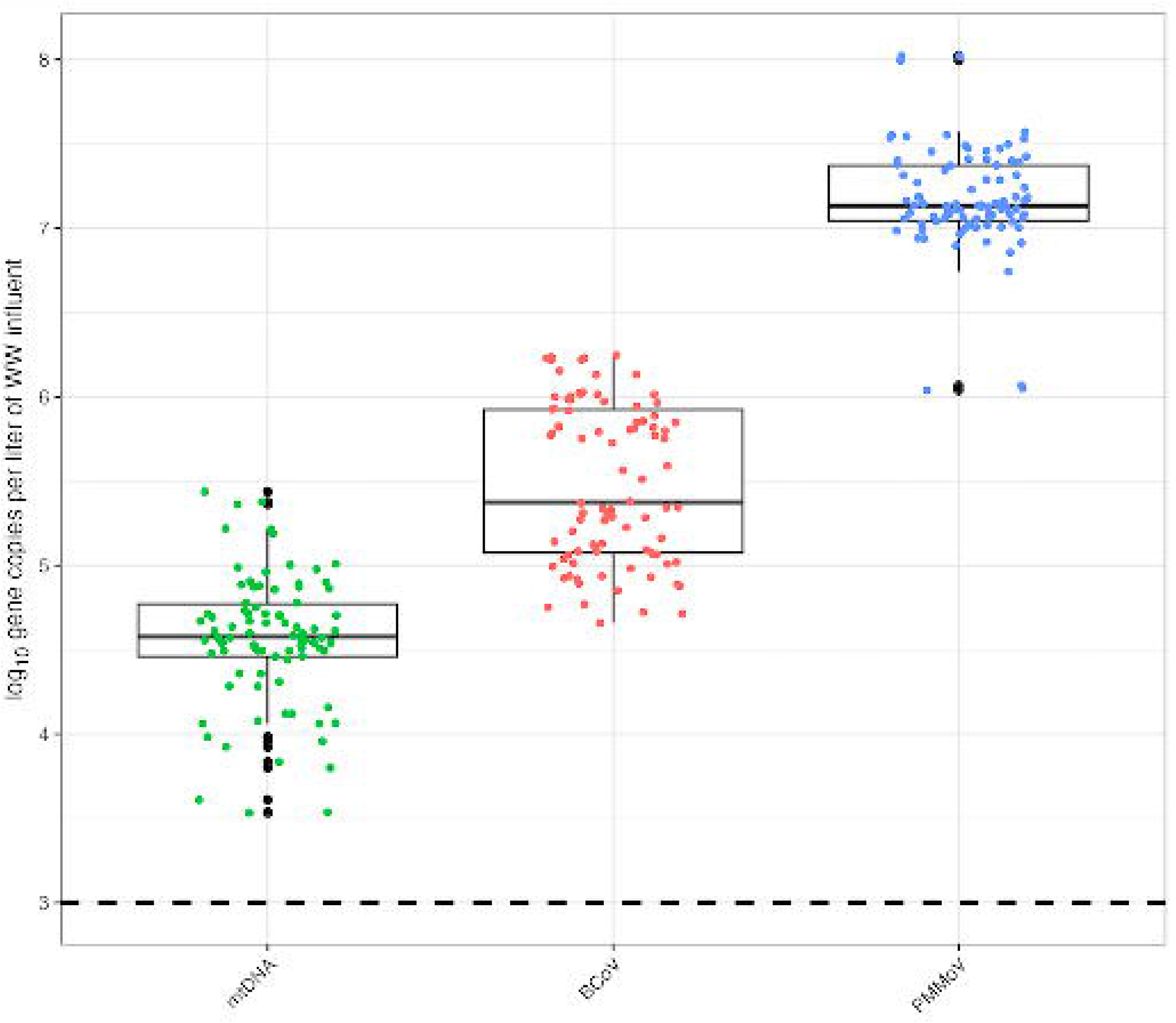
Log_10_ gene copies per liter of wastewater influent using the InnovaPrep Concentrating Pipette (CP) method. The dashed line represents the limit of detection when calculated as 3 partitions out of the total valid partitions. Figure includes all technical replicates per sample.

### Pathogen concentrations normalized by mtDNA and PMMoV

Quantitative log_10_ gene copies per liter of wastewater influent before (S9 Table) and after normalization (S10-11 Tables), with mtDNA normalization resulting in overall higher log_10_ ratios. In Fig 3, we note a considerably smaller ratio when using PMMoV normalization over mtDNA. These concentrations are caused by increased PMMoV concentrations in wastewater influent compared to mtDNA concentrations.

**Fig 3.**
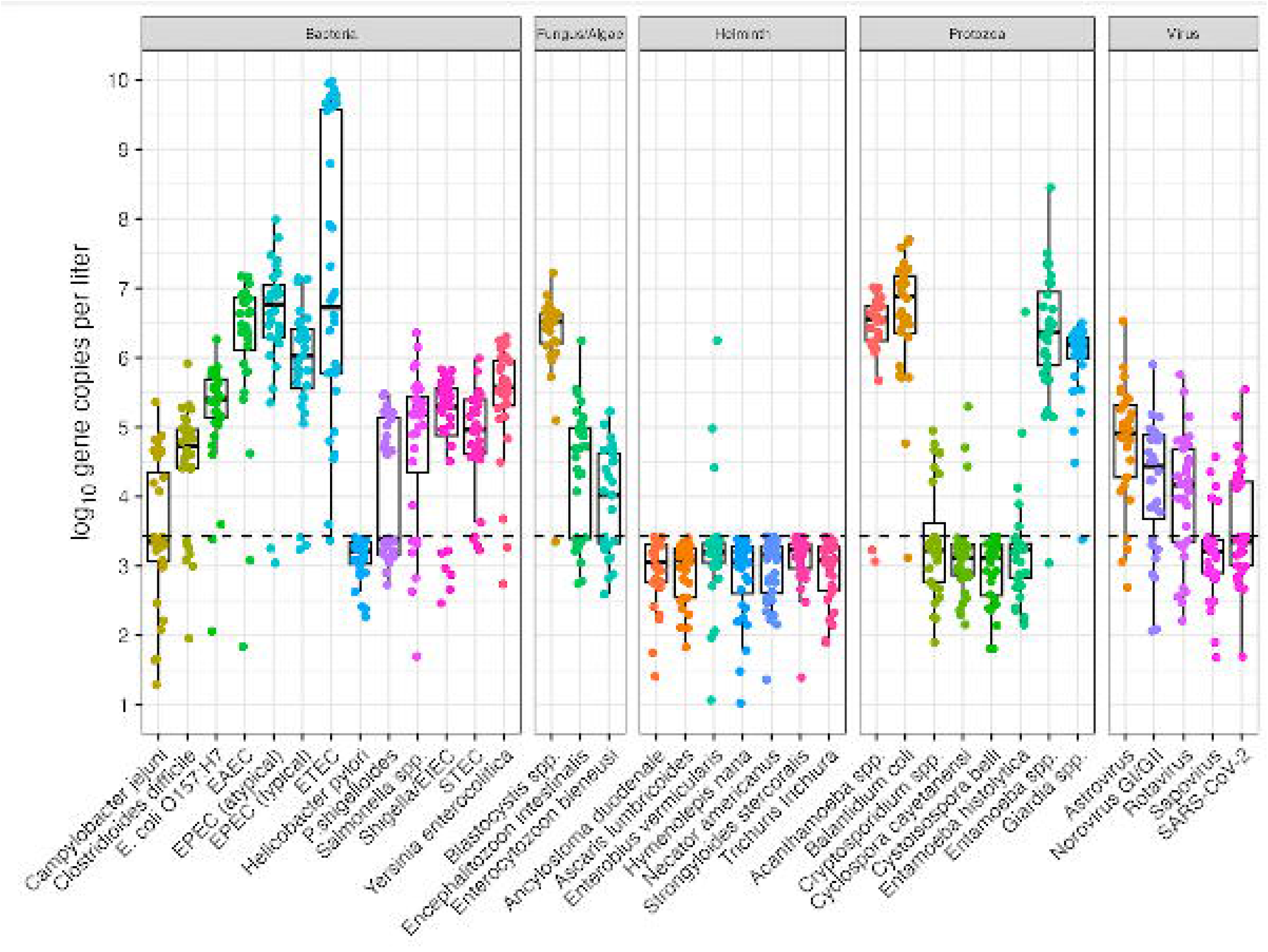
A) Pathogen data normalized by mtDNA. B) Pathogen data normalized by PMMoV. The dashed line represents where pathogen and normalizer count are equivalent. Figure includes all technical replicates per sample for mtDNA and PMMoV marker.

### TAC Performance Interpretation

#### Standard Curves

The standard curves for this custom TAC included two assays (Adenovirus 40/41 and Hepatitis A) with poor standard curve performances (r^2^ < .95) and therefore were excluded from all analyses. Of the remaining 40 enteric targets, the DNA control was phocine herpes virus and RNA control was MS2. For performance metrics (S12 Table), reasonable linearity was detected for all included assays with an average R^2^ value of 0.997 across all assays with the lowest R^2^ of 0.967 for STEC (stx2) and the highest R^2^ of 1 for *Acanthamoeba* spp., *Balantidium coli*, *E. coli* O157:H7, *Giardia* spp., *Plesiomonas shigelloides*, *Salmonella* spp., and STEC (stx1). The lowest efficiency assay was Astrovirus at 87% while the highest was *Entamoeba* spp. at 104%.

#### Effective Volume

The effective volume, which does not account for recovery efficiency, is calculated as the proportion of original wastewater sample assayed in a single qPCR reaction. The effective analyzed wastewater volume for InnovaPrep CP was 0.155 mL (SD 0.0605) per reaction and SMF was 0.410 mL of wastewater per reaction (SD 0.121).

#### Limit of Detection and matrix LOD

The 95% aLOD was calculated for each assay in S12 Table, reported as gene copies per reaction. The lowest detectable target as Cryptosporidium spp. at 0.6 gene copies per reaction and the highest as 291 gene copies per reaction for ETEC (LT), followed by 96 gene copies per reaction for STEC (stx2).

A comprehensive mLOD table for each assay indicates the gene copy per mL of sewage is found in S13 Table and includes the minimum, maximum, mean, standard deviations, standard error, and confidence intervals. These results indicate average gene copies per mL of wastewater influent as low as 1.591 for Cryptosporidium spp. and 16S marker or as high as 264.7 gc/mL for ETEC (LT & ST). SARS-CoV-2 mLOD was 16.4 gc/mL influent.

#### Inhibition

We used MS2 as the extraction control and the average Cq for negative extraction controls (n=7) was 17.8 gene copies per reaction [confidence interval 0.821], whereas all SMP samples (n=30) had an average Cq of 19.3 gene copies per reaction [CI 2.04]. With a Cq difference of 1.5, we can reasonably conclude inhibition was not a major issue with our sample matrix since samples and controls had Cq difference less than 2.

## Discussion

Wastewater surveillance sampling, processing, storage, and analysis methods have advanced rapidly since the emergence of SARS-CoV-2. Most studies have examined primary influent[34,35] and solids[36,37]. Sampling methods have also varied from grab, composite, and more recently passive techniques[38]. In addition to testing different matrices, many laboratories have implemented various methods to concentrate SARS-CoV-2 in wastewater using ultracentrifugation, polyethylene glycol precipitation, electronegative membrane filtration, and ultrafiltration[28,39], but few have considered a concentration step followed by a simultaneous, multi-parallel quantitative assay or multiple pathogen detection assays. The possibility of high-plex, high throughput platforms are of particular interest to stakeholders looking to expand wastewater monitoring nationally in the US and abroad. For example, the CDC has expanded upon the previously single-plex N1 assay for SARS-CoV-2 to include influenza A and/or B for increased testing capacity[40]. Practical applications of surveillance suggest that downstream sampling analyses of 3 or 4 samplings per week could provide useful results regarding trends, but the specific design would have to be driven by local public health trends and goals [41–43].

### TAC performance metrics

We compared our traditional metrics such as R^2^ trends of standard curves and found that our TAC results are within a reasonable R^2^ range for almost all assays (R^2^>0.96), except for two explicitly excluded due to poor standard curve performance. Our 95% LODs calculated also indicate a broad range of analytical sensitivities across all pathogen targets. With the lowest detections at 0.6 gene copies per reaction, we also have targets on the higher end of 291 gene copies per reaction for ETEC. While other studies indicate a loss of sensitivity when using TAC, there was still an 89% detection rate compared to single-plex assays run[44].

### Prevalence of bacteria, protozoan, and viral targets

Our qPCR data indicated 10^4^-10^6^ gene copies per liter for SARS-CoV-2 prior to normalization efforts, which is comparable to other studies [45]. Researchers had previously detected *Giardia duodenale*., *Cryptosporidium* spp., and *Enterocytozoon bieneusi* at 82.6%, 56.2% and 87.6%, in combined sewer overflows (CSO) around China[46]. These molecular surveillance findings were also similar to ours at 97% (n=29/30) for *Giardia* spp., not specifically *Giardia duodenale,* and 27% (n=8/30) for *Cryptosporidium* spp., and 53% (n=16/30) for *E. bieneusi*. Our data showed the presence of *Strongyloides stercoralis* in urban wastewater, a human parasite typically associated with rural, underserved settings[47]. This finding is an example of the utility of screening for uncommon or unexpected targets, revealing novel information that can supplement existing public health surveillance.

Groundwater and runoff can intrude into wastewater collection systems through inflow and infiltration (I&I), which may be relevant for fungi and a possibility for other microbial species to mix with wastewater flows[48]. Other potential explanations of sources into wastewater may include animal waste, commercial and/or industrial waste. These influent flows and their sources are difficult to determine, but routine surveillance – including with the addition of source-tracking – may provide additional insight into influent pathogens, their possible origins, and their utility in understanding infection transmission and control in the sewershed.

### Value of multiple detections on TAC

Multi-parallel detection of pathogens of interest using TAC can be helpful in long-term surveillance or monitoring of pathogens, including in rapid screening programs or where numerous pathogens may be of interest. Apart from known, emerging, or suspected pathogens, antimicrobial resistance genes or other PCR-detectable targets of public health relevance can be included in TAC design. One key premise of WBE and monitoring is the potential value of using the method as an early detection for the onset of a potential outbreak [49,50], yet most detection methods have a needle in a haystack approach versus a wider screening that could be especially applicable to state health departments or in routine monitoring. Most clinical testing is conducted one sample at a time and a high throughput method for simultaneous testing could expand the early warning potential to many other pathogens.

The customizability of TAC has proven useful in other applications such as surveillance of respiratory illness[51,52], acute-febrile illness for outbreak or surveillance purposes[53], and to improve etiological detection of difficult neonatal infectious diseases for low-resource clinical settings[54]. Some studies have focused on applications of combining nucleic acid detection with quantitative microbial risk assessments[55], but none have considered such a broad set of applications to wastewater monitoring and surveillance, although some have applied these methods qualitatively on fecal sludge samples[56,57]. It is possible to create a multiplex assay for digital PCR, the leading technology for wastewater monitoring, for up to five different genes, but no other platform provides quantitative data on up to 48 gene targets during a single experimental run.

TAC methods can fill a critical gap in existing molecular monitoring tools. As a method yielding quantitative estimates of potentially dozens of targets, it offers complementary advantages over emerging digital PCR platforms (greater sensitivity and lower limits of quantification, but fewer targets) and sequencing methods (many more targets, but high limits of detection and generally not quantitative). TAC should be considered where targets are present in high numbers – like in wastewaters and fecal sludges – and where many pathogens are of interest.

The application of improved methods for the detection and quantification of enteric pathogens in wastewater, in addition to other enteric pathogens of interest, can then be translated into relevant intervention and monitoring efforts[21]. As SARS-CoV-2 surveillance in wastewater reaches scale [7,34,58], detection and quantification of other pathogens has been proposed. Researchers have expanded on wastewater monitoring to focus increased surveillance on other respiratory viruses such as human influenza and rhinovirus[59], norovirus[60], or as an outbreak detection tool for influenza,[61] and are also considering other emerging infections such as monkeypox[62].

### Value of sensitivity of dPCR

The current and suggested methodology to process wastewater samples using a molecular platform is dPCR due to its low limit of detection and quantification. While these efforts make sense to consider when focused on one particular pathogen, it is not as feasible and consumes several resources if considering a truly practical monitoring system for wastewater. Time, technical staff labor and resources are always a challenge for laboratories and especially public health laboratories that have been tasked with monitoring wastewater for SARS-CoV-2. We can expect enteric targets to be present in wastewater, but to further identify which enteric pathogens are present and their concentrations with respect to each other would be a useful application towards building a wastewater monitoring system.

While SARS-CoV-2 was detected through TAC, we were also interested in detecting additionally relevant targets, including BCoV, PMMoV, and mtDNA, which were not previously included on the TAC. The normalization of pathogen concentrations using mtDNA consistently lowered concentrations across samples and may be useful as a normalization variable instead of or in addition to PMMoV. While PMMoV has been widely used for normalization of wastewater data[63,64], we found the normalization efforts did not drastically reduce the noise-to-signal ratio. While several studies have used PMMoV as a normalization marker for SARS-CoV-2[12,65,66], fewer studies have considered human mitochondrial DNA markers and those who have found the marker to have strong correlations to clinical case counts[67]. Additional studies have also considered the use of crAssphage[12,64], HF183[41,68], and *Bacteroides* ribosomal RNA (rRNA) and human 18S rRNA as other normalization markers to explore using for wastewater fecal concentration data[12]. Normalization techniques using a variety of biological (PMMoV, HF183, crAssphage) and chemical markers (ammonia, total kjeldahl nitrogen, total phosphorous, biochemical oxygen demand) have been proposed as a way of accounting for non-human inputs to sewers (i.e., dilution effects) and improving correlation with clinical data and comparability between sites. However, the effects of normalization with a variety of techniques on correlations with clinical data have been mixed [41,63,69–71]. Our observations are consistent with those of previous studies. Normalization with mtDNA nor PMMoV reduced the coefficient of variation for single analytes.

### Limitations

Wastewater sample recovery for SARS-CoV-2 has been successful when using fresh samples, but for many WWTP and their partners it may be unrealistic to complete same-day processing for logistical reasons[72]. This work demonstrated the recovery of pathogen targets using archived grab samples, which makes this approach open to a broader range of applications such as retrospective analyses where clinical data is available or can be linked to these environmental surveillance results. However, more research is needed to understand which recovery methods work best and can be performed efficiently for archived samples. While we did not optimize methods for recovery across all targets, it will be increasingly important to consider such methods when screening for multiple targets and depending on target selection [68,73,74].

A major limitation to interpreting this work is limited data on using multiple TAC targets and their incorporation into predictive models. Researchers have gained interest in calculating community-specific or dorm-specific fecal shedding rates specifically for SARS-CoV-2[75,76], but there was no specific information on the fecal shedding rates for this particular population to consider a modeling approach to relate pathogen concentration and clinical case data for asymptomatic individuals. Additionally, sewersheds of different sizes may have specific challenges in determining accurate shedding rates. Robust data on enteric shedding rates is not widely available for high-income countries, but efforts to estimate these variables and their uncertainties have been attempted[77].

TAC methods are also limited by the number of gene target detections one can consider. With the option of detecting many pathogens comes with a need for determining the most relevant genes of interest. While TAC can run up to 48 unique targets, the total amount of template that enters each individual well is ∼ 0.6 μL. This low template volume, compared to a 2-5 μL of template included in other molecular assays can affect the overall limits of detection for this platform. While singleplex assays may have lower limits of detection, the likelihood of optimizing a multiplex for up to 46 or more agents is unrealistic; therefore, giving TAC a considerable advantage as a high parallel, multiple detection platform[44].

Additionally, these targets and the QA/QC involved require dedicated time and effort to include relevant targets that may change based on future applications. The need for additional replicates run to produce robust analytical limits of quantification are encouraged for future work. Using this multiple pathogen detection tool does not account for variant changes and may not be suitable for all applications. Our findings indicate TAC offers a multi-parallel platform for screening wastewater for a diverse array of enteric pathogens of interest to public health with strong potential for screening other targets of interest including respiratory viruses and antibiotic resistance genes.

## Supporting information

Supplemental References & Legends

## Data Availability

All data from this manuscript have been posted in a publicly-accessible database and are available at this link: https://osf.io/rg36f/

## Acknowledgements

The authors would like to acknowledge the wastewater treatment plants that permitted sample collection and the National Science Foundation (NSF 2027752-Collaborative Research-RAPID: Wastewater Informed Epidemiological Monitoring of SARS-CoV-2) for financial support in completing this work. DC and AL were supported in part by T32ES007018, Biostatistics for Research in Environmental Health, funded by the National Institute of Environmental Health Sciences to the University of North Carolina at Chapel Hill. The funders had no role in study design, data collection and analysis, decision to publish, or preparation of the manuscript. GR led the initial draft, GR and DC led data analysis, JB and AB conceived the work, and the following authors worked on data collection and sample processing: KZ, AK, YL, RC, AL, JK, GR. All authors reviewed and edited the manuscript.

