## Supplemental Table 1 for "Simultaneous detection and quantification of multiple pathogen targets in wastewater"

**S1 Table.** pH, temperature and Total Suspended Solids (TSS) for all wastewater influent (n=30)

|  | Total Suspended Solids (mg/L) | pH | Temperature (°C) |
| --- | --- | --- | --- |
| Mean (SD) | 194.7 (125.8) | 7.15 (0.52) | 7.15 (1.28) |
| Min | 0 | 6.0 | 4.5 |
| Max | 640 | 8.1 | 9.0 |
| Median | 162 | 7.20 | 7.4 |
| IQR | 139 | 0.54 | 2.1 |
