## Supplemental Table 8 for "Simultaneous detection and quantification of multiple pathogen targets in wastewater"

**S8 Table.** BCoV percent recovery by sample

| Sample # | Recovery % |
| --- | --- |
| 1 | 23.7 |
| 2 | 36 |
| 3 | 38.3 |
| 4 | 48.1 |
| 5 | 77.8 |
| 6 | 56.3 |
| 7 | 57.5 |
| 8 | 58.6 |
| 9 | 4.99 |
| 10 | 95.1 |
| 11 | 32.4 |
| 12 | 50.9 |
| 13 | 35.1 |
| 14 | 96.7 |
| 15 | 41.2 |
| 16 | 11 |
| 17 | 6.45 |
| 18 | 7.31 |
| 19 | 8.12 |
| 20 | 4.49 |
| 21 | 2.82 |
| 22 | 5.33 |
| 23 | 10.2 |
| 24 | 5.03 |
| 25 | 3.5 |
| 26 | 6.95 |
| 27 | 10.9 |
| 28 | 11.5 |
| 29 | 20.3 |
| 30 | 12.6 |
