## Supplemental Table 7 for "Simultaneous detection and quantification of multiple pathogen targets in wastewater"

**S7 Table.** dMIQE Checklist

| **ITEM TO CHECK** | **PROVIDED** | **COMMENT** |
| --- | --- | --- |
| **Column1** | **Y/N** | **Column2** |
| **1. SPECIMEN** |  |  |
| Detailed description of specimen type and numbers | **Y** | See Methods section: Sample Collection |
| Sampling procedure (including time to storage) | **Y** | See Methods section: Sample Collection |
| Sample aliquotation, storage conditions and duration | **Y** | See Methods section: Sample Collection |
| **2. NUCLEIC ACID EXTRACTION** |  |  |
| Description of extraction method including amount of sample processed | **Y** | See Methods section |
| Volume of solvent used to elute/resuspend extract | **Y** | 100 µL to elute |
| Number of extraction replicates | **N** | Not tested |
| Extraction blanks included? | **N** | Not tested |
| **3. NUCLEIC ACID ASSESSMENT AND STORAGE** |  |  |
| Method to evaluate quality of nucleic acids |  |  |
| Method to evaluate quantity of nucleic acids (including molecular weight and calculations when using mass) | **Y** | Qubit 4 Fluorometer |
| Storage conditions: temperature, concentration, duration, buffer, aliquots | **Y** | Text S1 |
| Clear description of dilution steps used to prepare working DNA solution | **Y** | No dilutions run |
| **4. NUCLEIC ACID MODIFICATION** |  |  |
| Template modification (digestion, sonication, pre-amplification, bisulphite etc.) | **N** | No template modification |
| Details of repurification following modification if performed | **N** | No repurification performed |
| **5. REVERSE TRANSCRIPTION** |  |  |
| cDNA priming method and concentration | **Y** | Did not conduct |
| One or two step protocol (include reaction details for two step) | **Y** | One-Step |
| Amount of RNA added per reaction | **Y** | 5 µL |
| Detailed reaction components and conditions | **Y** | See Text S1 |
| Estimated copies measured with and without addition of RT* | **N** | Did not conduct |
| Manufacturer of reagents used with catalogue and lot numbers | **Y** | Qiagen, QIAcuity One-Step RT-PCR Kit |
| Storage of cDNA: temperature, concentration, duration, buffer and aliquots | **Y** | See Text S1 |
| **6. dPCR OLIGONUCLEOTIDES DESIGN AND TARGET INFORMATION** |  |  |
| Sequence accession number or official gene symbol | **Y** | hCYTB484, BCoV, PMMoV |
| Method (software) used for design and *in silico* verification | **N** | Used literature references to obtain primers/probes (Table S4) |
| Location of amplicon | **Y** | Text S2 |
| Amplicon length | **Y** | Text S2 |
| Primer and probe sequences (or amplicon context sequence)** | **Y** | See Table S1 |
| Location and identity of any modifications | **Y** | No modifications performed |
| Manufacturer of oligonucleotides | **Y** | Integrated DNA Technologies, Inc. (IDT) |
| **7. dPCR PROTOCOL** |  |  |
| Manufacturer of dPCR instrument and instrument model | **Y** | Qiagen, QIAcuity Four |
| Buffer/kit manufacturer with catalogue and lot number | **Y** | QIAcuity One-Step Viral RT-PCR Kit, Catalogue Number: 1123145 |
| Primer and probe concentration | **Y** | See Table S5 |
| Pre-reaction volume and composition (incl. amount of template and if restriction enzyme added) | **Y** | 5 µL, no restriction enzyme added |
| Template treatment (initial heating or chemical denaturation) | **N** | No template treatment |
| Polymerase identity and concentration, Mg++ and dNTP concentrations*** | **N** | Proprietary; Qiagen, Hilden, Germany |
| Complete thermocycling parameters | **Y** | See Text S1 |
| **8. ASSAY VALIDATION** |  |  |
| Details of optimisation performed | **Y** | See Text S1 |
| Analytical specificity (vs. related sequences) and limit of blank (LOB) | **Y** | LOB = NTC |
| Analytical sensitivity/LoD and how this was evaluated | **N** | No additional sensitivity tests run |
| Testing for inhibitors (from biological matrix/extraction) | **N** | No additional inhibition tests run (see Results section: Inhibition) |
| **9. DATA ANALYSIS** |  |  |
| Description of dPCR experimental design |  |  |
| Comprehensive details negative and positive of controls (whether applied for QC or for estimation of error) | **Y** | See Text S1 |
| Partition classification method (thresholding) | **Y** | See Text S1 |
| Examples of positive and negative experimental results (including fluorescence plots in supplemental material) | **Y** | See Figure S1 |
| Description of technical replication | **Y** | See Text S2 |
| Repeatability (intra-experiment variation) | **Y** | Technical replicates run in triplicate |
| Reproducibility (inter-experiment/user/lab etc. variation ) | **N** | No other lab/user variation |
| Number of partitions measured (average and standard deviation ) | **Y** | See Text S2 |
| Partition volume | **N** | Calculated by software |
| Copies per partition (λ or equivalent ) (average and standard deviation) | **Y** | BCoV= 0.03 (sd=0.03); PMMoV = 1.3 (sd=1.2); mtDNA = 3.4x10^-3^ (sd= 3.1x10^-3^) |
| dPCR analysis program (source, version) | **Y** | See Text S1 |
| Description of normalisation method | **Y** | We normalized gene copy estimates |
| Statistical methods used for analysis | **Y** | See Methods section: Sample Analysis |
| Data transparency | Available at OSF | <https://osf.io/rg36f/> |
