## Supplemental Table 13 for "Simultaneous detection and quantification of multiple pathogen targets in wastewater"

**S13 Table.** 95% Matrix Limit of Detection (gene copies per mL sewage)

| Target | Min | Max | Mean | Standard Deviation | Standard Error | Confidence Interval |
| --- | --- | --- | --- | --- | --- | --- |
| 16S | 0.86 | 3.05 | 1.59 | 0.526 | 0.096 | 0.196 |
| *Acanthamoeba* spp. | 32.95 | 116.9 | 60.99 | 20.15 | 3.679 | 7.524 |
| *Ancylostoma duodenale* | 8.88 | 31.53 | 16.44 | 5.432 | 0.992 | 2.028 |
| *Ascaris lumbricoides* | 8.88 | 31.53 | 16.44 | 5.432 | 0.992 | 2.028 |
| astrovirus | 8.88 | 31.53 | 16.44 | 5.432 | 0.992 | 2.028 |
| *Blantidium coli* | 3.15 | 11.19 | 5.83 | 1.927 | 0.352 | 0.72 |
| *Blastocystis* spp. | 3.15 | 11.19 | 5.83 | 1.927 | 0.352 | 0.72 |
| *Campylobacter jejuni/coli* | 30.09 | 106.8 | 55.69 | 18.398 | 3.359 | 6.87 |
| *Clostridium difficile* | 8.88 | 31.53 | 16.44 | 5.432 | 0.992 | 2.028 |
| *Cryptosporidium* spp. | 0.86 | 3.05 | 1.59 | 0.526 | 0.096 | 0.196 |
| *Cyclospora cayetanensi* | 3.15 | 11.19 | 5.83 | 1.927 | 0.352 | 0.72 |
| *Cystoisospora belli* | 8.88 | 31.53 | 16.44 | 5.432 | 0.992 | 2.028 |
| *E. coli* O157:H7 | 3.15 | 11.19 | 5.83 | 1.927 | 0.352 | 0.72 |
| *Entamoeba histolytica* | 8.88 | 31.53 | 16.44 | 5.432 | 0.992 | 2.028 |
| EAEC (aatA and aaiC) | 8.88 | 116.98 | 38.72 | 26.81 | 3.461 | 6.926 |
| EPEC (eae & bfpa) | 3.15 | 31.53 | 11.14 | 6.704 | 0.865 | 1.732 |
| ETEC (LT & ST) | 3.15 | 1480.1 | 264.7 | 388.9 | 40.99 | 81.44 |
| *Encephalitozoon intestinalis* | 3.15 | 11.19 | 5.83 | 1.927 | 0.352 | 0.72 |
| *Entamoeba* spp. | 30.09 | 106.8 | 55.69 | 18.398 | 3.359 | 6.87 |
| *Enterobius vermicularis* | 103.2 | 366.2 | 190.9 | 63.08 | 11.52 | 23.55 |
| *Enterocytozoon bieneusi* | 6.88 | 24.41 | 12.73 | 4.205 | 0.768 | 1.57 |
| *Giardia* spp. | 8.88 | 31.53 | 16.44 | 5.432 | 0.992 | 2.028 |
| *Helicobacter pylori* | 8.88 | 31.53 | 16.44 | 5.432 | 0.992 | 2.028 |
| *Hymenolepis nana* | 3.15 | 11.19 | 5.83 | 1.927 | 0.352 | 0.72 |
| MS2 | 1.43 | 5.086 | 2.65 | 0.876 | 0.16 | 0.327 |
| *Necator americanus* | 6.16 | 21.87 | 11.40 | 3.767 | 0.688 | 1.407 |
| norovirus GI/GII | 32.95 | 116.98 | 60.99 | 19.98 | 2.579 | 5.161 |
| PHhpv | 8.88 | 31.53 | 16.44 | 5.432 | 0.992 | 2.028 |
| *Plesiomonas shigelloides* | 32.95 | 116.98 | 60.99 | 20.15 | 3.68 | 7.52 |
| rotavirus | 8.88 | 31.53 | 16.44 | 5.432 | 0.992 | 2.028 |
| SARS-CoV-2 | 8.88 | 31.53 | 16.44 | 5.432 | 0.992 | 2.028 |
| STEC (stx1 & stx2) | 103.2 | 488.3 | 222.8 | 80.39 | 10.38 | 20.77 |
| *Salmonella* spp. | 3.15 | 11.19 | 5.83 | 1.927 | 0.352 | 0.72 |
| sapovirus I/II/IV/V | 3.15 | 11.19 | 5.83 | 1.911 | 0.247 | 0.494 |
| *Strongyloides stercoralis* | 3.15 | 11.19 | 5.83 | 1.927 | 0.352 | 0.72 |
| *Trichuris trichiura* | 3.15 | 11.19 | 5.83 | 1.927 | 0.352 | 0.72 |
| *Yersinia enterocolitica* | 3.15 | 11.19 | 5.83 | 1.927 | 0.352 | 0.72 |
| *Shigella*/EIEC | 32.95 | 116.98 | 60.99 | 20.15 | 3.679 | 7.524 |
