## Supplemental Table 12 for "Simultaneous detection and quantification of multiple pathogen targets in wastewater"

**S12 Table.** TAC performance and 95% LOD

| Target | Target Gene | slope | y-intercept | R^2^ | Efficiency | 95% limit of detection† |
| --- | --- | --- | --- | --- | --- | --- |
| enteric 16S | 16S | -3.31 | 38.9 | 0.998 | 101% | 0.60 |
| *Acanthamoeba* spp. | 18S rRNA | -3.39 | 37.8 | 1.000 | 97% | 23 |
| adenovirus 40/41* | Fiber gene | NA | NA | 0.670 | NA | NA |
| *Ancylostoma duodenale* | ITS-2 | -3.38 | 39.1 | 1.000 | 98% | 6.2 |
| *Ascaris lumbricoides* | ITS-1 | -3.45 | 38.6 | 1.000 | 95% | 6.2 |
| astrovirus | Capsid | -3.69 | 37.5 | 0.998 | 87% | 6.2 |
| *Balantidium coli* | ITS-1 | -3.39 | 37.9 | 1.000 | 97% | 2.2 |
| *Blastocystis* spp. | 18S rRNA | -3.32 | 40.6 | 0.997 | 100% | 2.2 |
| *Cystoisospora belli* | 18S rRNA | -3.35 | 37.8 | 0.999 | 99% | 6.2 |
| *Cyclospora cayetanensi* | 18S rRNA | -3.34 | 37.2 | 0.998 | 99% | 2.2 |
| *Campylobacter jejuni/coli* | *cadF* | -3.34 | 38.3 | 0.999 | 99% | 21 |
| *Clostridioides difficile* | *tcdB* | -3.43 | 37.5 | 0.999 | 96% | 6.2 |
| *Cryptosporidium* spp. | 18S rRNA | -3.40 | 38.0 | 0.999 | 97% | 0.6 |
| DNA control (phocine herpes virus) | *gB* | -3.32 | 37.0 | 0.998 | 100% | 6.2 |
| *Enterocytozoon bieneusi* | ITS | -3.28 | 37.2 | 0.999 | 102% | 4.8 |
| *E. coli* O157:H7 | *rfbE* | -3.46 | 38.0 | 1.000 | 95% | 2.2 |
| *Encephalitozoon intestinalis* | SSU rRNA | -3.38 | 38.5 | 0.999 | 98% | 2.2 |
| *Enterobius vermicularis* | 5S | -3.46 | 38.6 | 0.999 | 95% | 72 |
| EAEC (aaiC) | *aaiC* | -3.42 | 38.2 | 0.999 | 96% | 6.2 |
| EAEC (aatA) | *aatA* | -3.43 | 37.7 | 0.998 | 96% | 23 |
| *Entamoeba histolytica* | 18S rRNA | -3.28 | 38.0 | 0.996 | 102% | 6.2 |
| *Entamoeba* spp. | 18S rRNA | -3.23 | 37.3 | 0.974 | 104% | 21 |
| EPEC (typical) | *bfpA* | -3.38 | 37.5 | 0.999 | 98% | 6.2 |
| EPEC (atypical) | *eae* | -3.37 | 37.6 | 0.999 | 98% | 2.2 |
| ETEC (LT) | *LT* | -3.46 | 47.6 | 0.990 | 94% | 291 |
| ETEC (STh) | *STh* | -3.38 | 38.8 | 0.999 | 98% | 6.2 |
| ETEC (STp) | *STp* | -3.35 | 37.3 | 0.999 | 99% | 2.2 |
| *Giardia* spp. | 18S rRNA | -3.42 | 37.9 | 1.000 | 96% | 6.2 |
| *Hymenolepis nana* | ITS-1 | -3.38 | 38.2 | 1.000 | 98% | 2.2 |
| *Helicobacter pylori* | *ureC* | -3.41 | 37.7 | 0.998 | 97% | 6.2 |
| hepatitis A virus* | NCR | -2.73 | NA | 0.840 | 132% | NA |
| *Shigella*/EIEC | *ipaH* | -3.35 | 37.5 | 0.999 | 99% | 23 |
| MS2 (RNA control) | *MS2g1* | -3.58 | 37.5 | 0.999 | 90% | 1.0 |
| *Necator americanus* | ITS-2 | -3.37 | 39.8 | 1.000 | 98% | 4.3 |
| norovirus GII | ORF1-2 | -3.54 | 37.0 | 0.999 | 92% | 23 |
| norovirus GI | ORF1-2 | -3.49 | 35.9 | 0.997 | 93% | 23 |
| *Plesiomonas shigelloides* | *gyrB* | -3.42 | 38.2 | 1.000 | 96% | 23 |
| rotavirus | NSP3 | -3.57 | 38.0 | 0.998 | 91% | 6.2 |
| *Salmonella* spp. | *invA* | -3.42 | 38.4 | 1.000 | 96% | 2.2 |
| sapovirus I/II/IV | RdRp | -3.66 | 38.2 | 0.998 | 88% | 2.2 |
| sapovirus V | RdRp | -3.56 | 36.7 | 0.999 | 91% | 2.2 |
| SARS-CoV-2 | N1 | -3.53 | 36.2 | 0.995 | 92% | 6.2 |
| *Strongyloides stercoralis* | Dispersed repetitive sequence | -3.33 | 37.5 | 0.999 | 100% | 2.2 |
| STEC (stx1) | *stx1* | -3.41 | 39.9 | 1.000 | 97% | 72 |
| STEC (stx2) | *stx2* | -3.37 | 38.3 | 0.967 | 98% | 96 |
| *Trichuris trichiura* | 18S rRNA | -3.35 | 38.4 | 1.000 | 99% | 2.2 |
| *Yersinia enterocolitica* | *lytA* | -3.48 | 38.3 | 0.998 | 94% | 2.2 |

*Excluded due to poor standard curve performance

†Stokdyk *et al*. 2016[8]; units are gene copies per reaction
