## Supplemental Table 11 for "Simultaneous detection and quantification of multiple pathogen targets in wastewater"

**S11 Table.** Normalization using PMMoV— mean (standard deviation) - excluding duplicates

| Target | WWTP A | WWTP B | WWTP C | WWTP D |
| --- | --- | --- | --- | --- |
| *Acanthamoeba* spp. | -0.9 (0.3) | -0.3 (0.7) | -0.5 (0.3) | -1.0 (0.1) |
| adenovirus | NA | NA | NA | NA |
| *Ancylostoma duodenale* | -- | -- | -- | -- |
| *Ascaris lumbricoides* | -- | -- | -- | -- |
| astrovirus | -2.6 (0.4) | -2.0 (1.3) | -2.1 (0.6) | -2.3 (0) |
| *Balantidium coli* | -1.3 (0.3) | -0.8 (0.5) | -0.1 (0.5) | -1.7 (0.1) |
| *Blastocystis* spp. | -1.6 (0.3) | -0.9 (0) | -0.6 (0.3) | -0.9 (0.2) |
| *Campylobacter jejuni/coli* | -- | -3.9 (0) | -2.6 (0.5) | -- |
| *Clostridioides difficile* | -2.6 (0.3) | -1.8 (0.4) | -2.3 (0.4) | -1.9 (0.4) |
| *Cryptosporidium* spp. | -- | -- | -2.5 (0.3) | -- |
| *Cyclospora cayetanensi* | -- | -- | -2.6 (0.4) | -- |
| *Cystoisospora belli* | -- | -- | -- | -- |
| *E. coli* O157:H7 | -2.0 (0.2) | -2.4 (0.9) | -1.7 (0.4) | -1.6 (0.4) |
| EAEC* | -0.7 (0.1) | -1.7 (0.3) | -0.5 (0.5) | -0.4 (0) |
| EPEC (atypical)† | -0.6 (0.2) | -1.3 (0.5) | -0.4 (0.5) | 0.1 (0.4) |
| EPEC (typical)† | -1.1 (0.3) | -1.9 (0.2) | -1.1 (0.4) | -0.4 (0) |
| ETEC* | 2.0 (0.1) | 0.9 (0.5) | 2.5 (0.5) | 3.0 (0.3) |
| *Encephalitozoon intestinalis* | -2.9 (0.4) | -2.4 (0) | -2.0 (0.5) | -2.4 (0.2) |
| *Entamoeba histolytica* | -- | -- | -3.4 (0.8) | -- |
| *Entamoeba* spp. | -1.8 (0.3) | -0.9 (0.1) | -0.4 (0.7) | -2.0 (0.3) |
| *Enterobius vermicularis* | -- | -- | -1.8 (0.8) | -- |
| *Enterocytozoon bieneusi* | -2.8 (0) | -3.3 (0) | -2.7 (0.4) | -2.8 (0) |
| *Giardia* spp. | -2.1 (0.7) | -1.3 (0.3) | -1.0 (0.3) | -1.3 (0.1) |
| *Helicobacter pylori* | -- | -- | -- | -- |
| hepatitis A | NA | NA | NA | NA |
| *Hymenolepis nana* | -- | -- | -- | -- |
| *Necator americanus* | -- | -- | -- | -- |
| norovirus GI/GII* | -3.3 (0.7) | -1.2 (0) | -2.7 (0.7) | -2.8 (0.5) |
| *Plesiomonas shigelloides* | -2.9 (0.1) | -- | -1.9 (0.3) | -2.8 (0) |
| rotavirus | -2.7 (0.2) | -2.1 (0) | -2.6 (0.6) | -3.4 (0.5) |
| SARS-CoV-2 | -3.6 (0.3) | -3.3 (0) | -2.4 (0.5) | -- |
| STEC* | -2.2 (0.2) | -1.9 (0) | -1.6 (0.5) | -1.4 (0.1) |
| *Salmonella* spp. | -2.2 (0.3) | -1.8 (0) | -2.0 (0.5) | -1.3 (0.3) |
| sapovirus* | -- | -- | -2.8 (0.1) | -- |
| *Shigella*/EIEC† | -2.5 (0.1) | -- | -1.7 (0.4) | -2.4 (0.6) |
| *Strongyloides stercoralis* | -- | -- | -- | -- |
| *Trichuris trichiura* | -- | -- | -- | -- |
| *Yersinia enterocolitica* | -2.1 (0.1) | -2.2 (0.6) | -1.4 (0.4) | -2.1 (0) |

*Enteroaggregative *E. coli* (EAEC) combined gene targets aatA and aaiC; enterotoxigenic *E. coli* (ETEC) combined targets from gene LT, STh, and STp; norovirus included GI and GII targets; sapovirus combined gene targets for I, II, IV, and V; shiga toxin-producing *E. coli* (STEC) combined gene targets stx1 and stx2.

†Enteropathogenic *E. coli* (EPEC); enteroinvasive *E. coli* (EIEC)
