## Supplemental Table 10 for "Simultaneous detection and quantification of multiple pathogen targets in wastewater"

**S10 Table.** Normalization using mtDNA – mean (standard deviation) - excluding duplicates

| Target | WWTP A | WWTP B | WWTP C | WWTP D |
| --- | --- | --- | --- | --- |
| *Acanthamoeba* spp. | 1.9 (0.1) | 2.5 (0.7) | 2.1 (0.3) | 1.1 (0.2) |
| adenovirus | NA | NA | NA | NA |
| *Ancylostoma duodenale* | -- | -- | -- | -- |
| *Ascaris lumbricoides* | -- | -- | -- | -- |
| astrovirus | 0.1 (0.5) | 0.9 (1.0) | 0.5 (0.6) | -- |
| *Balantidium coli* | 1.5 (0.1) | 2.0 (0.2) | 2.5 (0.4) | 0.5 (0.2) |
| *Blastocystis* spp. | 1.1 (0.1) | 1.8 (0.3) | 2.0 (0.3) | 1.3 (0.3) |
| *Campylobacter jejuni/coli* | -- | -1.3 (0) | 0.1 (0.6) | -- |
| *Clostridioides difficile* | 0.1 (0.1) | 1.0 (0.1) | 0.3 (0.3) | 0.3 (0.5) |
| *Cryptosporidium* spp. | -- | -- | 0 (0.2) | -- |
| *Cyclospora cayetanensi* | -- | -- | 0.4 (0.4) | -- |
| *Cystoisospora belli* | -- | -- | -- | -- |
| *E. coli* O157:H7 | 0.7 (0.2) | 0.3 (1.2) | 0.9 (0.5) | 0.6 (0.4) |
| EAEC* | 2.1 (0.2) | 1.0 (0.5) | 2.1 (0.5) | 1.8 (0.1) |
| EPEC (atypical)† | 2.1 (0.1) | 1.6 (0.2) | 2.2 (0.5) | 2.3 (0.5) |
| EPEC (typical)† | 1.7 (0.5) | 1.0 (0.2) | 1.6 (0.5) | 1.8 (0.1) |
| ETEC* | 4.8 (0.2) | 3.6 (0.8) | 5.1 (0.5) | 5.2 (0.3) |
| *Encephalitozoon intestinalis* | -0.1 (0.4) | 0.7 (0.1) | 0.6 (0.5) | -0.2 (0.3) |
| *Entamoeba histolytica* | -- | -- | -0.7 (0.8) | -- |
| *Entamoeba* spp. | 0.8 (0.1) | 1.7 (0.4) | 2.1 (0.7) | 0.1 (0.4) |
| *Enterobius vermicularis* | -- | -- | 1 (0.9) | -- |
| *Enterocytozoon bieneusi* | 0.1 (0.3) | -0.1 (0.1) | 0 (0.4) | -0.7 (0) |
| *Giardia* spp. | 0.6 (0.5) | 1.4 (0.4) | 1.7 (0.3) | 0.9 (0.2) |
| *Helicobacter pylori* | -- | -- | -- | -- |
| hepatitis A | NA | NA | NA | NA |
| *Hymenolepis nana* | -- | -- | -- | -- |
| *Necator americanus* | -- | -- | -- | -- |
| norovirus GI/GII* | -0.5 (0.9) | 1.3 (0) | 0 (0.6) | -0.6 (0.5) |
| *Plesiomonas shigelloides* | 0 (0.2) | -- | 0.7 (0.3) | -0.7 (0) |
| rotavirus | 0.2 (0.5) | 0.4 (0) | 0 (0.5) | -1.2 (0.6) |
| SARS-CoV-2 | -0.8 (0) | -0.8 (0) | 0.2 (0.4) | -- |
| STEC* | 0.6 (0.4) | 1.0 (0.3) | 1.0 (0.6) | 0.8 (0) |
| *Salmonella* spp. | 0.5 (0.2) | 0.8 (0) | 0.7 (0.5) | 0.8 (0.4) |
| sapovirus* | -- | -- | -0.4 (0.3) | -- |
| *Shigella*/EIEC† | 0.4 (0.2) | -- | 0.9 (0.4) | -0.2 (0.5) |
| *Strongyloides stercoralis* | -- | -- | -- | -- |
| *Trichuris trichiura* |  | -- | -- | -- |
| *Yersinia enterocolitica* | 0.7 (0.4) | 0.7 (0.9) | 1.3 (0.4) | 0 (0.1) |

*Enteroaggregative *E. coli* (EAEC) combined gene targets aatA and aaiC; enterotoxigenic *E. coli* (ETEC) combined targets from gene LT, STh, and STp; norovirus included GI and GII targets; sapovirus combined gene targets for I, II, IV, and V; shiga toxin-producing *E. coli* (STEC) combined gene targets stx1 and stx2.

†Enteropathogenic *E. coli* (EPEC); enteroinvasive *E. coli* (EIEC)
