## Supplemental Table 9 for "Simultaneous detection and quantification of multiple pathogen targets in wastewater"

**S9 Table.** Mean log_10_ gene copy concentrations per liter of WW influent (standard deviation) before normalization, by WW Treatment plant – skim milk flocculation

| Target | WWTP A | WWTP B | WWTP C | WWTP D |
| --- | --- | --- | --- | --- |
| *Acanthamoeba* spp. | 6.8 (0.1) | 4.9 (2.9) | 6.2 (1.5) | 6.4 (0.1) |
| adenovirus | NA | NA | NA | NA |
| *Ancylostoma duodenale* | 0 (0) | 0 (0) | 0 (0) | 0 (0) |
| *Ascaris lumbricoides* | 0 (0) | 0 (0) | 0 (0) | 0 (0) |
| astrovirus | 5.0 (0.5) | 2.3 (2.4) | 4.7 (1.3) | 2.7 (2.6) |
| *Balantidium coli* | 6.4 (0.1) | 4.5 (2.6) | 7.0 (0.4) | 5.8 (0.2) |
| *Blastocystis* spp. | 6.0 (0.1) | 4.4 (2.6) | 6.5 (0.3) | 6.6 (0.1) |
| *Campylobacter jejuni/coli* | 0 (0) | 0.8 (1.4) | 2.7 (2.2) | 0 (0) |
| *Clostridioides difficile* | 5.0 (0.2) | 2.4 (2.4) | 4.0 (1.8) | 5.5 (0.4) |
| *Cryptosporidium* spp. | 0 (0) | 0 (0) | 2.0 (2.3) | 0 (0) |
| *Cyclospora cayetanensi* | 0 (0) | 0 (0) | 0.8 (1.8) | 0 (0) |
| *Cystoisospora belli* | 0 (0) | 0 (0) | 0 (0) | 0 (0) |
| *E. coli* O157:H7 | 5.6 (0.2) | 3.6 (2.2) | 5.3 (0.5) | 5.8 (0.4) |
| EAEC* | 7.0 (0.2) | 3.8 (2.2) | 6.6 (0.5) | 7.1 (0.1) |
| EPEC (atypical)† | 7.0 (0.1) | 2.9 (3.0) | 6.7 (0.5) | 7.6 (0.4) |
| EPEC (typical)† | 6.6 (0.4) | 2.6 (2.6) | 6.0 (0.4) | 7.1 (0.2) |
| ETEC* | 9.7 (0.2) | 6.0 (3.5) | 9.6 (0.4) | 10.5 (0.2) |
| *Encephalitozoon intestinalis* | 4.8 (0.4) | 1.2 (2.1) | 3.4 (2.4) | 5.0 (0.2) |
| *Entamoeba histolytica* | 0 (0) | 0 (0) | 1.2 (1.8) | 0 (0) |
| *Entamoeba* spp. | 5.9 (0.2) | 4.4 (2.6) | 6.7 (0.7) | 5.5 (0.3) |
| *Enterobius vermicularis* | 0 (0) | 0 (0) | 0.6 (1.7) | 0 (0) |
| *Enterocytozoon bieneusi* | 3.3 (2.4) | 1.0 (1.7) | 2.9 (2.1) | 2.3 (2.4) |
| *Giardia* spp. | 5.5 (0.5) | 4.1 (2.5) | 6.1 (0.3) | 6.2 (0.1) |
| *Helicobacter pylori* | 0 (0) | 0 (0) | 0 (0) | 0 (0) |
| hepatitis A | NA | NA | NA | NA |
| *Hymenolepis nana* | 0 (0) | 0 (0) | 0 (0) | 0 (0) |
| *Necator americanus* | 0 (0) | 0 (0) | 0 (0) | 0 (0) |
| norovirus GI/GII* | 4.4 (0.9) | 1.2 (2.1) | 4.0 (1.5) | 4.7 (0.4) |
| *Plesiomonas shigelloides* | 3.3 (2.3) | 0 (0) | 2.8 (2.6) | 2.4 (2.4) |
| rotavirus | 3.4 (2.5) | 1.0 (1.7) | 3.3 (2.1) | 4.1 (0.5) |
| SARS-CoV-2 | 2.8 (2.0) | 0.7 (1.2) | 1.8 (2.3) | 0.2 (0.8) |
| STEC* | 5.5 (0.4) | 2.6 (2.6) | 5.1 (1.4) | 6.1 (0.2) |
| *Salmonella* spp. | 5.4 (0.2) | 1.3 (2.4) | 4.8 (1.3) | 5.9 (1.2) |
| *sapovirus** | 0 (0) | 0 (0) | 0.5 (1.4) | 0.2 (0.9) |
| *Shigella*/EIEC† | 3.5 (2.5) | 0 (0) | 5.1 (1.3) | 5.1 (0.6) |
| *Strongyloides stercoralis* | 0 (0) | 0 (0) | 0 (0) | 0 (0) |
| *Trichuris trichiura* | 0 (0) | 0 (0) | 0 (0) | 0 (0) |
| *Yersinia enterocolitica* | 5.6 (0.3) | 2.5 (2.5) | 5.7 (0.4) | 5.3 (0.3) |

*Enteroaggregative *E. coli* (EAEC) combined gene targets aatA and aaiC; enterotoxigenic *E. coli* (ETEC) combined targets from gene LT, STh, and STp; norovirus included GI and GII targets; sapovirus combined gene targets for I, II, IV, and V; shiga toxin-producing *E. coli* (STEC) combined gene targets stx1 and stx2.

†Enteropathogenic *E. coli* (EPEC); enteroinvasive *E. coli* (EIEC)
