## Supplemental Table 6 for "Simultaneous detection and quantification of multiple pathogen targets in wastewater"

**S6 Table.** Prevalence of pathogens detected in wastewater influent from four treatment plants in Atlanta, Georgia – using InnovaPrep concentrating pipette pellet

| Target | WW Plant A  (n=3) | WW Plant B  (n=3) | WW Plant C  (n=18) | WW Plant D  (n=4) |
| --- | --- | --- | --- | --- |
| *Acanthamoeba* spp. | 3 (100%) | 3 (100%) | 18 (100%) | 4 (100%) |
| adenovirus | NA | NA | NA | NA |
| *Ancylostoma duodenale* | 0 (-) | 0 (-) | 0 (-) | 0 (-) |
| *Ascaris lumbricoides* | 0 (-) | 0 (-) | 0 (-) | 0 (-) |
| astrovirus | 0 (-) | 0 (-) | 3 (17%) | 0 (-) |
| *Balantidium coli* | 3 (100%) | 3 (100%) | 18 (100%) | 4 (100%) |
| *Blastocystis* spp. | 3 (100%) | 3 (100%) | 17 (94%) | 4 (100%) |
| *Campylobacter jejuni* | 0 (-) | 3 (100%) | 0 (-) | 0 (-) |
| *Clostridioides difficile* | 3 (100%) | 3 (100%) | 10 (56%) | 1 (25%) |
| *Cryptosporidium* spp. | 0 (-) | 1 (33%) | 3 (17%) | 0 (-) |
| *Cyclospora cayetanensi* | 0 (-) | 0 (-) | 2 (11%) | 0 (-) |
| *Cystoisospora belli* | 0 (-) | 0 (-) | 0 (-) | 0 (-) |
| *E. coli* O157:H7 | 2 (67%) | 3 (100%) | 7 (39%) | 2 (50%) |
| EAEC* | 3 (100%) | 3 (100%) | 18 (100%) | 4 (100%) |
| *Encephalitozoon intestinalis* | 3 (100%) | 3 (100%) | 15 (83%) | 4 (100%) |
| *Entamoeba histolytica* | 0 (-) | 0 (-) | 2 (11%) | 0 (-) |
| *Entamoeba* spp. | 3 (100%) | 3 (100%) | 14 (78%) | 3 (75%) |
| *Enterobius vermicularis* | 0 (-) | 0 (-) | 2 (11%) | 0 (-) |
| *Enterocytozoon bieneusi* | 3 (100%) | 2 (67%) | 11 (61%) | 1 (25%) |
| EPEC (atypical)† | 3 (100%) | 3 (100%) | 18 (100%) | 4 (100%) |
| EPEC (typical)† | 3 (100%) | 3 (100%) | 17 (94%) | 4 (100%) |
| ETEC* | 3 (100%) | 3 (100%) | 18 (100%) | 4 (100%) |
| *Giardia* spp. | 2 (67%) | 3 (100%) | 18 (100%) | 4 (100%) |
| *Helicobacter pylori* | 0 (-) | 0 (-) | 1 (6%) | 0 (-) |
| hepatitis A | NA | NA | NA | NA |
| *Hymenolepis nana* | 0 (-) | 0 (-) | 3 (17%) | 0 (-) |
| *Necator americanus* | 0 (-) | 0 (-) | 0 (-) | 0 (-) |
| norovirus GI/GII* | 0 (-) | 0 (-) | 6 (33%) | 1 (25%) |
| *Plesiomonas shigelloides* | 0 (-) | 0 (-) | 2 (11%) | 0 (-) |
| rotavirus | 0 (-) | 0 (-) | 3 (17%) | 0 (-) |
| *Salmonella* spp. | 2 (67%) | 2 (67%) | 3 (17%) | 0 (-) |
| sapovirus* | 0 (-) | 0 (-) | 6 (33%) | 2 (50%) |
| SARS-CoV-2 | 0 (-) | 0 (-) | 3 (17%) | 0 (-) |
| *Shigella*/EIEC† | 2 (67%) | 0 (-) | 13 (72%) | 3 (75%) |
| STEC* | 3 (100%) | 0 (-) | 7 (39%) | 4 (100%) |
| *Strongyloides stercoralis* | 0 (-) | 0 (-) | 0 (-) | 1 (25%) |
| *Trichuris trichiura* | 0 (-) | 0 (-) | 0 (-) | 0 (-) |
| *Yersinia enterocolitica* | 2 (67%) | 3 (100%) | 10 (56%) | 0 (-) |

*Enteroaggregative *E. coli* (EAEC) combined gene targets aatA and aaiC; enterotoxigenic *E. coli* (ETEC) combined targets from gene LT, STh, and STp; norovirus included GI and GII targets; sapovirus combined gene targets for I, II, IV, and V; shiga toxin-producing *E. coli* (STEC) combined gene targets stx1 and stx2.

†Enteropathogenic *E. coli* (EPEC); enteroinvasive *E. coli* (EIEC)
