## Supplemental Table 5 for "Simultaneous detection and quantification of multiple pathogen targets in wastewater"

**S5 Table.** dPCR Primer and Probe Sequences

| **Target Name** | **Description** | **Oligonucleotide Sequence (5'>3')** | **Detection Channel** | **Label** | **Concentrations** |
| --- | --- | --- | --- | --- | --- |
| N1 | 2019-nCoV_N1 Forward Primer | GAC CCC AAA ATC AGC GAA AT | Green | -- | 400 nM |
|  | 2019-nCoV_N1 Reverse Primer | TCT GGT TAC TGC CAG TTG AAT CTG |  | -- | 400 nM |
|  | 2019-nCoV_N1 Probe | ACC CCG CAT TAC GTT TGG TGG ACC |  | FAM, ZEN, Iowa Black FQ | 200 nM |
| mtDNA | hCYTB484 Forward Primer | CAATGAATCTGAGGAGGCTAC | Yellow | -- | 200 nM |
|  | hCYTB484 Reverse Primer | CGTGCAAGAATAGGAGGTG |  | -- | 200 nM |
|  | hCYTB520TM Probe | ACCCTCACACGATTCTTTACCTTTCACT |  | SUN, Iowa Black RQ | 100 nM |
| BCoV | BCoV Forward Primer | CTGGAAGTTGGTGGAGTT | Orange | -- | 3200 nM |
|  | BCoV Reverse Primer | ATTATCGGCCTAACATACATC |  | -- | 3200 nM |
|  | BCoV Probe | CCTTCATATCTATACACATCAAGTTGTT |  | TAMRA, Iowa Black RQ | 400 nM |
| PMMoV | PMMoV Forward Primer | GAGTGGTTTGACCTTAACGTTGA | Red | -- | 3200 nM |
|  | PMMoV Reverse Primer | TTGTCGGTTGCAATGCAAGT |  | -- | 3200 nM |
|  | PMMoV Probe | CCTACCGAAGCAAATG |  | Texas Red, Iowa Black RQ | 400 nM |
