## Supplemental Table 4 for "Simultaneous detection and quantification of multiple pathogen targets in wastewater"

**S4 Table.** MIQE Checklist

| **ITEM TO CHECK** | **IMPORTANCE** | **CHECKLIST** |
| --- | --- | --- |
| **EXPERIMENTAL DESIGN** |  |  |
| Definition of experimental and control groups | **E** | Cross-sectional study with no experimental or control group |
| Number within each group | **E** | 30 wastewater samples were analyzed |
| Assay carried out by core lab or investigator's lab? | D | Investigator's lab |
| Acknowledgement of authors' contributions | D | See Acknowledgements section |
| **SAMPLE** |  |  |
| Description | **E** | See Methods Section |
| Volume/mass of sample processed | D | 750 mL wastewater influent; 200 mg pellet input for extraction |
| Microdissection or macrodissection | **E** | Not applicable |
| Processing procedure | **E** | Frozen at -80°C |
| If frozen - how and how quickly? | **E** | Same day as collection |
| If fixed - with what, how quickly? | **E** | Not fixed |
| Sample storage conditions and duration (especially for FFPE samples) | **E** | Frozen at -80°C for ~1 year |
| **NUCLEIC ACID EXTRACTION** |  |  |
| Procedure and/or instrumentation | **E** | See Methods Section |
| Name of kit and details of any modifications | **E** | Powersoil Pro - Manual extraction |
| Source of additional reagents used | D | Not applicable |
| Details of DNase or RNAse treatment | **E** | Not applicable |
| Contamination assessment (DNA or RNA) | **E** | One extraction control was included for each extraction day |
| Nucleic acid quantification | **E** | Qubit 1x HS dsDNA kit |
| Instrument and method | **E** | Qubit 4 Fluorometer |
| Purity (A260/A280) | D | Not measured |
| Yield | D | Not measured |
| RNA integrity method/instrument | **E** | Not measured |
| RIN/RQI or Cq of 3' and 5' transcripts | **E** | Not measured |
| Electrophoresis traces | D | Not applicable |
| Inhibition testing (Cq dilutions, spike or other) | **E** | Monitored amplification of inhibition control |
| **REVERSE TRANSCRIPTION** |  |  |
| Complete reaction conditions | **E** | One-step reverse transcription |
| Amount of RNA and reaction volume | **E** | Reaction Volume = 1.5 µL; Template volume = 0.6 µL/well |
| Priming oligonucleotide (if using GSP) and concentration | **E** | Proprietary |
| Reverse transcriptase and concentration | **E** | ArrayScript Reverse Transcriptase |
| Temperature and time | **E** | 45°C for 20 minutes |
| Manufacturer of reagents and catalogue numbers | D | Applied Biosystems, Ag-Path-ID One-Step RT-PCR Reagents, Catalog number: 4387391 |
| Cqs with and without RT | D* | Not measured |
| Storage conditions of cDNA | D | Not applicable |
| **qPCR TARGET INFORMATION** |  |  |
| If multiplex, efficiency and LOD of each assay. | **E** | S12 Table |
| Sequence accession number | **E** | NA |
| Location of amplicon | D | S2 Text |
| Amplicon length | **E** | S2 Text |
| *In silico* specificity screen (BLAST, etc) | **E** | All assays were BLASTed to confirm specificity before ordering the custom TAC |
| Pseudogenes, retropseudogenes or other homologs? | D | Not applicable |
| Sequence alignment | D | Not applicable |
| Secondary structure analysis of amplicon | D | Not applicable |
| Location of each primer by exon or intron (if applicable) | **E** | Not applicable |
| What splice variants are targeted? | **E** | Not applicable |
| **qPCR OLIGONUCLEOTIDES** |  |  |
| Primer sequences | **E** | S3 Table |
| RTPrimerDB Identification Number | D | Not available |
| Probe sequences | D** | S3 Table |
| Location and identity of any modifications | **E** | No modifications |
| Manufacturer of oligonucleotides | D | ThermoFisher Scientific |
| Purification method | D | Not applicable |
| **qPCR PROTOCOL** |  |  |
| Complete reaction conditions | **E** | 45°C for 20 min and 95°C for 10 min, followed by 45 cycles of 95°C for 15 s and 60°C for 1 min |
| Reaction volume and amount of cDNA/DNA | **E** | 38 µL of template with 62 µL of AgPath-ID™ One-Step RT-PCR Reagents |
| Primer, (probe), Mg++ and dNTP concentrations | **E** | All assays contained the same concentrations of primers (900 nanomolar) and probe (250 nanomolar). The Mg2+ and dNTP concentrations are not listed in the in the User Guide. |
| Polymerase identity and concentration | **E** | AmpliTaq Gold™ polymerase |
| Buffer/kit identity and manufacturer | **E** | AgPath-ID™ One-Step RT-PCR Reagents |
| Exact chemical constitution of the buffer | D | Proprietary |
| Additives (SYBR Green I, DMSO, etc.) | **E** | No additives |
| Manufacturer of plates/tubes and catalog number | D | ThermoFisher Scientific |
| Complete thermocycling parameters | **E** | 45°C for 20 min and 95°C for 10 min, followed by 45 cycles of 95°C for 15 s and 60°C for 1 min |
| Reaction setup (manual/robotic) | D | Manual set-up in a disinfected dead air box (10% bleach with fifteen minutes of contact time, UV for fifteen minutes, and a final cleaning step with 70% ethanol) |
| Manufacturer of qPCR instrument | **E** | ThermoFisher Scientific |
| **qPCR VALIDATION** |  |  |
| Evidence of optimisation (from gradients) | D | S12 Table |
| Specificity (gel, sequence, melt, or digest) | **E** | Not applicable |
| For SYBR Green I, Cq of the NTC | **E** | Not applicable |
| Standard curves with slope and y-intercept | **E** | S12 Table |
| PCR efficiency calculated from slope | **E** | S12 Table |
| Confidence interval for PCR efficiency or standard error | D | Not available |
| r2 of standard curve | **E** | S12 Table |
| Linear dynamic range | **E** | Not provided |
| Cq variation at lower limit | **E** | Not provided |
| Confidence intervals throughout range | D | Not provided |
| Evidence for limit of detection | **E** | S12 Table |
| If multiplex, efficiency and LOD of each assay. | **E** | Technically, not multiplex |
| **DATA ANALYSIS** |  |  |
| qPCR analysis program (source, version) | **E** | Design and Analysis Real-Time PCR Software (version 2.6.0) |
| Cq method determination | **E** | Manual thresholding; Cq cut-off = 40 |
| Outlier identification and disposition | **E** | Not applicable |
| Results of NTCs | **E** | We observed no amplification in our 5 PCR negative template controls, except for the 16S assay, MS2 extraction control, PhHV extraction control, and internal manufacturer control (S5 Fig). |
| Justification of number and choice of reference genes | **E** | Not applicable |
| Description of normalisation method | **E** | Normalized to mtDNA and PMMoV markers |
| Number and concordance of biological replicates | D | See Results Section |
| Number and stage (RT or qPCR) of technical replicates | **E** | See Results Section |
| Repeatability (intra-assay variation) | E | Duplicates performed |
| Reproducibility (inter-assay variation, %CV) | D | Not available |
| Power analysis | D | Not applicable |
| Statistical methods for result significance | **E** | See Methods Section |
| Software (source, version) | E | R Studio v4.2.1 & Design & Analysis 2.6.0 |
| Cq or raw data submission using RDML | **D** | Data available upon request |
