## Supplemental References & Legends for "Simultaneous detection and quantification of multiple pathogen targets in wastewater"

List of Legends

**S1 Table.** pH, temperature and Total Suspended Solids (TSS) for all wastewater influent (n=30)

**S2 Table. Matched samples comparison on TAC for pathogen types**

**S3 Table.** qPCR Primer and Probe Sequences for TAC

**S4 Table.** MIQE Checklist

**S5 Table.** dPCR Primer and Probe Sequences

**S6 Table.** Prevalence of pathogens detected in wastewater influent from four treatment plants in Atlanta, Georgia – using InnovaPrep concentrating pipette pellet

**S7 Table.** dMIQE Checklist

**S8 Table.** BCoV percent recovery by sample

**S9 Table.** Mean log_10_ gene copy concentrations per liter of WW influent (standard deviation) before normalization, by WW Treatment plant – skim milk flocculation

**S10 Table.** Normalization using mtDNA – mean (standard deviation) - excluding duplicates

**S11 Table.** Normalization using PMMoV— mean (standard deviation) - excluding duplicates

**S12 Table.** TAC performance and 95% LOD

**S13 Table.** 95% Matrix Limit of Detection (gene copies per mL sewage)

**S1 Text.** Multiplex assay optimization (BCoV, PMMoV, N1, mtDNA)

**S2 Text.** dPCR assay details, including positive and negative control results

**S1 Fig.** BCoV, PMMoV, mtDNA dPCR RFU plots displaying threshold partitioning for samples, positive and no template controls (NTC)

**S2 Fig.** InnovaPrep Concentrating Pipette Pellet TAC Results

**S3 Fig.** Direct extraction TAC boxplot

**S4 Fig.** Skim Milk Flocculation TAC Boxplot Results by gene target

**S5 Fig.** Amplification and multicomponent plot for no-template control. The amplification occurs for MS2, PhHV, manufacture internal control, and 16S.
