## Supplemental Figure 5 for "Simultaneous detection and quantification of multiple pathogen targets in wastewater"

**S5 Fig.** Amplification and multicomponent plot for no-template control. The amplification occurs for MS2, PhHV, manufacture internal control, and 16S.

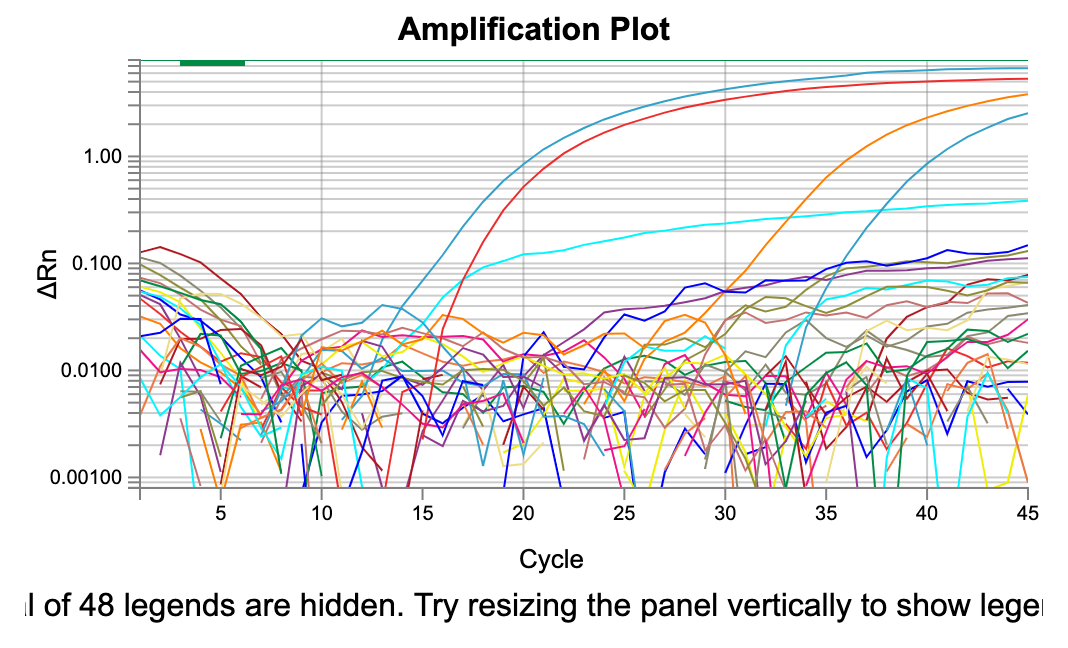

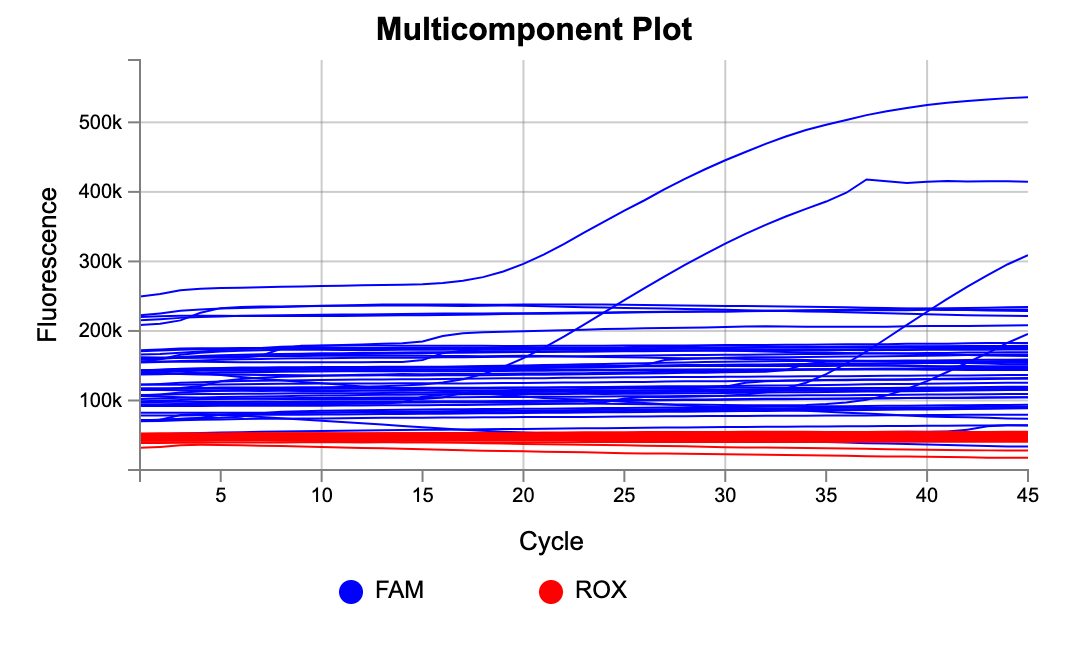
