## Supplemental Figure 4 for "Simultaneous detection and quantification of multiple pathogen targets in wastewater"

**S4 Fig.** Skim Milk Flocculation TAC Boxplot Results by gene target

Boxplot for Skim Milk Flocculation – DNeasy PowerSoil Pro Manual extractions (n=30). The dashed line represents the log_10_-transformed theoretical limit of detection (1 gene copy per reaction).

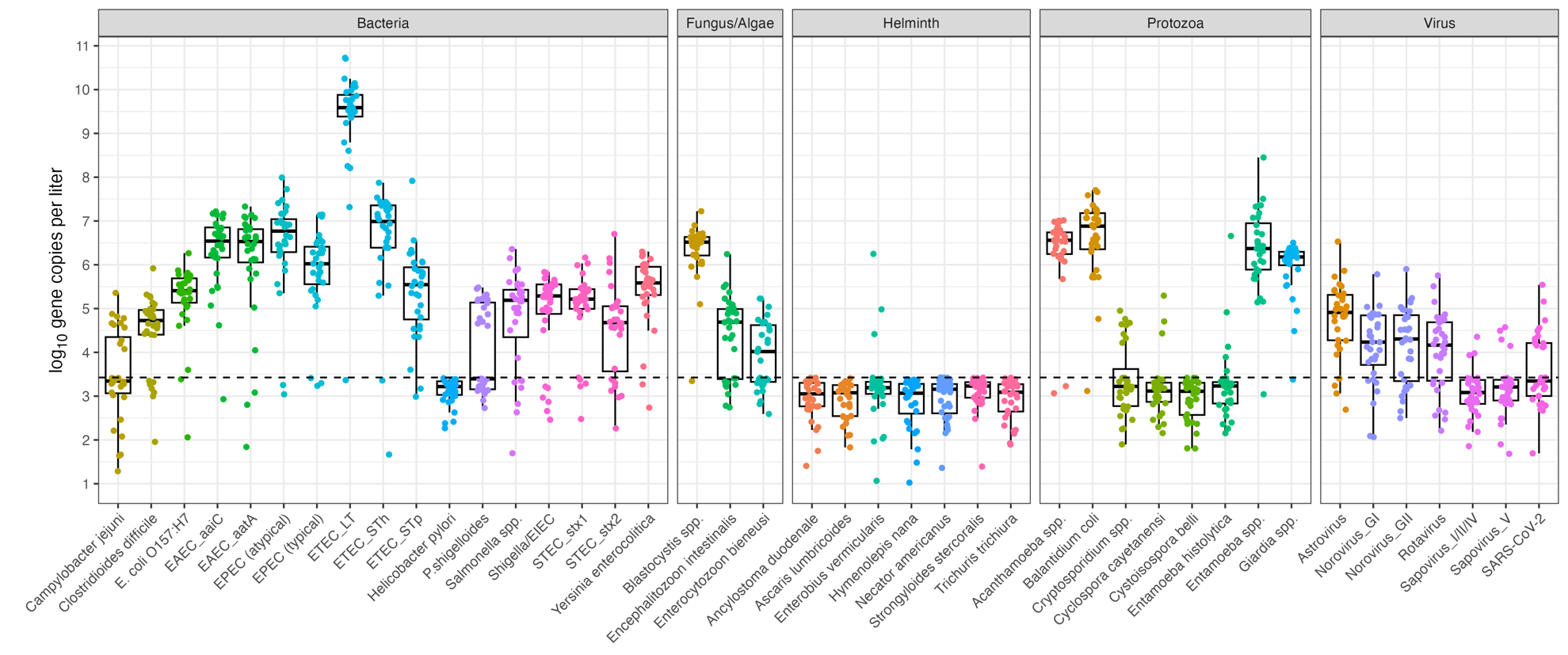
