## Supplemental Figure 3 for "Simultaneous detection and quantification of multiple pathogen targets in wastewater"

**S3 Fig.** Direct extraction TAC boxplot

Boxplot for direct extractions – DNeasy PowerSoil Pro Manual extractions (n= 4). The dashed line represents the log_10_-transformed theoretical limit of detection (1 gene copy per reaction).

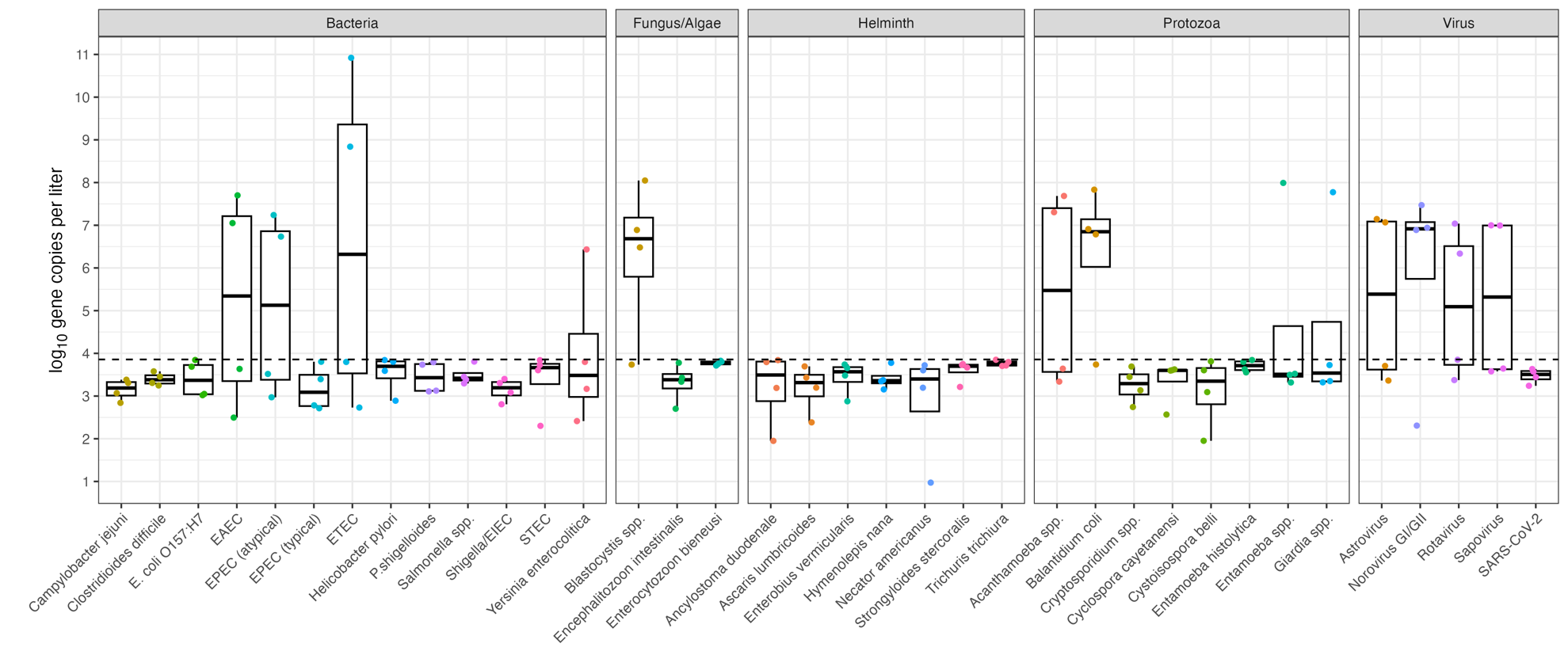
