## Supplemental Figure 2 for "Simultaneous detection and quantification of multiple pathogen targets in wastewater"

**S2 Fig.** InnovaPrep Concentrating Pipette Pellet TAC Results

Boxplot for InnovaPrep Concentrating Pipette Pellet – DNeasy PowerSoil Pro Manual extractions (n= 28 [N=30, 2 duplicates included in plots below]). The dashed line represents the log_10_-transformed theoretical limit of detection (1 gene copy per reaction).

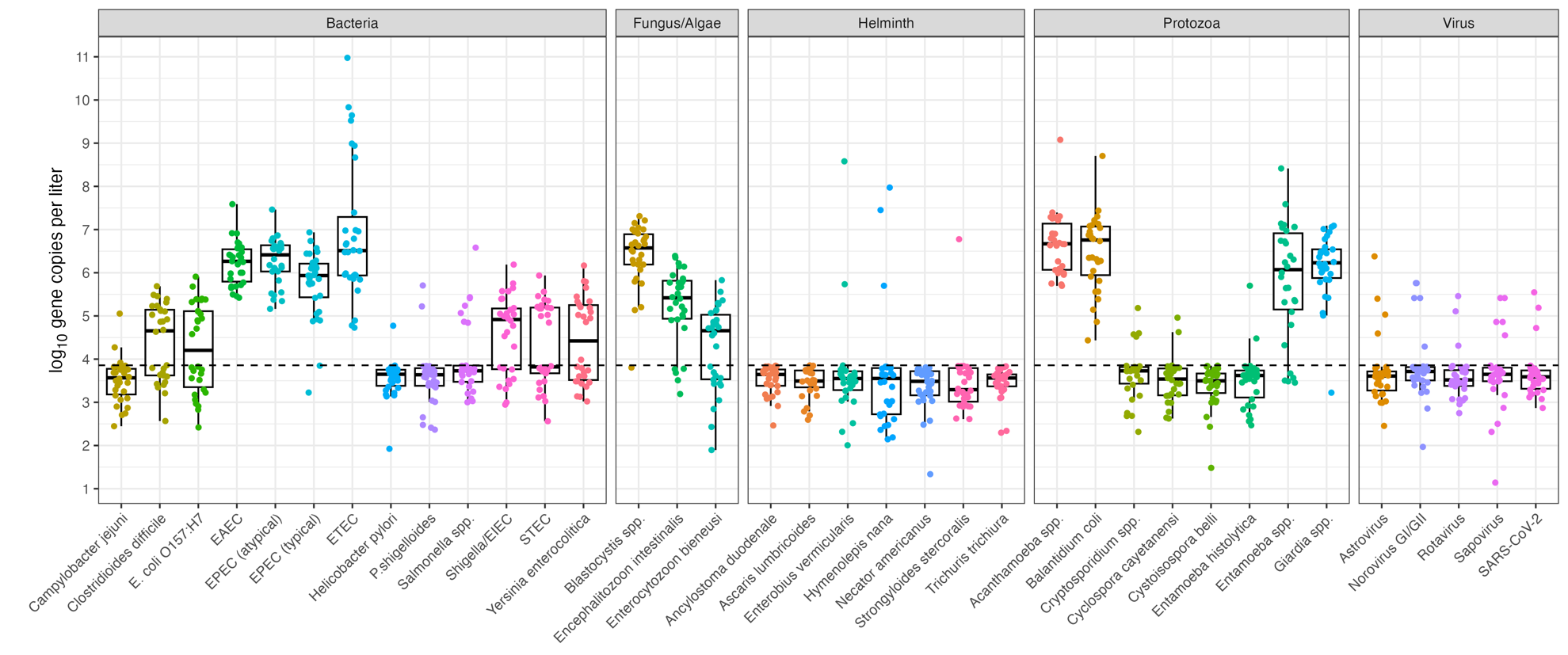

We processed an average of 139 mL through the InnovaPrep CP and the average CP pellet was 1.2 mL.
