## Supplemental Figure 1 for "Simultaneous detection and quantification of multiple pathogen targets in wastewater"

**S1 Fig.** BCoV, PMMoV, mtDNA dPCR RFU plots displaying threshold partitioning for samples, positive and no template controls (NTC)
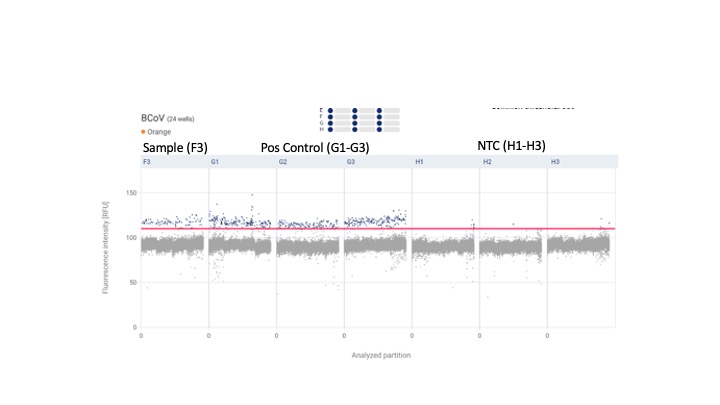

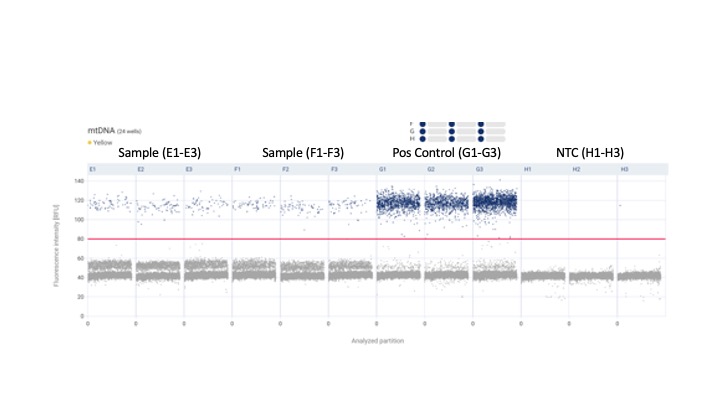

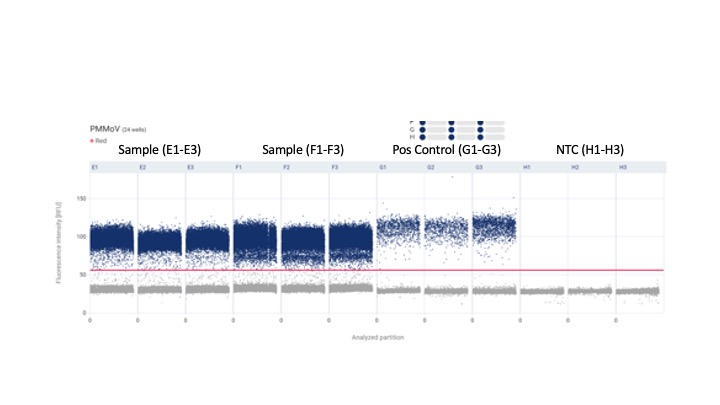
