## Supplemental Text 2 for "Simultaneous detection and quantification of multiple pathogen targets in wastewater"

**S2 Text.** dPCR assay details, including positive and negative control results

We procured gBlocks (Integrated DNA Technologies [IDT], Coralville, Iowa) for positive control (PC) templates and no template controls (NTC) were conducted with molecular grade water. We used Calf-Guard Vaccine (Zoetis) as the BCoV positive control. The Calf-Guard vaccine was resuspended with 2 mL of RNase/DNase-free water using a sterile 1 mL syringe. Aliquots of 41 μL were placed into a low-bind 1.5 mL microcentrifuge tube and placed in the -20°C freezer until use.

We included one NTC and one PC in triplicate for each batch of plates run and analyzed sample extracts over four days. We observed below detection limits for the three NTCs performed and positive controls had the following average copies per μL: mtDNA= 63.9 copies/μL, N1= 81.6 copies/μL, PMMoV = 70.9 copies/μl, BCoV = 147.7 copies/μL. Target amplicons and amplicon lengths are included below.

The average number of valid partitions per reaction for BCoV was 25298 (standard deviation [SD] = 242), N1 was 25309 (SD=225), PMMoV was 25286 (SD=261), and mtDNA was 25308 (SD=226).

| Amplicon Target | Amplicon Sequence (5’ > 3’) | Nucleotide bases (nts) |
| --- | --- | --- |
| N1 | GACCCCAAAATCAGCGAAATGCACCCCGCATTACGTTTGGTGGACCCTCAGATTCAACTGGCAGTAACCAGA | 72 nts |
| BCoV | CTGGAAGTTGGTGGAGTTTCAACCCAGAAACAAACAACTTGATGTGTATAGATATGAAGGGAAGGATGTATGTTAGGCCGATAAT | 85 nts |
| PMMoV | GAGTGGTTTGACCTTAACGTTTGAGCGGCCTACCGAAGCAAATGTCGCACTTGCATTGCAACCGACAA | 68 nts |
| hCYTB484 | CAATGAATCTGAGGAGGCTACTCAGTAGACAGTCCCACCCTCACACGATTCTTTACCTTTCACTTCATCTTACCCTTCATTATTGCAGCCCTAGCAGCACTCCACCTCCTATTCTTGCACG | 121 nts |
