## Supplemental Text 1 for "Simultaneous detection and quantification of multiple pathogen targets in wastewater"

**S1 Text.** Multiplex assay optimization (BCoV, PMMoV, N1, mtDNA)

Digital PCR assays were run as single-plex to determine optimal primer/probe concentrations, cycling conditions, and were then combined to determine cross-channel harmony (Table 2). All final sample extracts were run using Qiagen One-step Viral RT-PCR Mastermix (260 μL) along with the multiplex RT mix (10.4 μL), the combined primers and probes (170.8 μL, BCoV = 121.7 μL), template (5 μL) and the remaining volume replaced with nuclease-free water until 45 μL/reaction well was reached. The QIAcuity Nanoplate 26k 24-well plates were used for each sample run in triplicate and sealed with a Nanoplate roller before placing into the QIAcuity Four. The QIAcuity Four machine used the QIAcuity Software Suite 2.1.7.182 (Qiagen, Hilden, Germany). A positive and negative control was run for each batch. While we attempted to design a four-plex, issues with the orange channel fluorescence and cross-over from other channels suggested to single-plex the orange channel target (bovine coronavirus (BCoV)). All final cycling conditions include 50°C at 40 minutes, one cycle at 95°C for 2 minutes, and 40 cycles at 95°C for 5 seconds and 50°C for 1 minute. The red, yellow, and green channels were imaged together, and the orange channel was imaged separately with reduced exposure by 100 m/s. Positive control samples were used for thresholding positive and negative partition bands with the following cut-offs mtDNA: 80 RFU (fluorescence intensity), N1: 40 RFU, PMMoV: 56 RFU, BCoV: 110 RFU.

Assay optimization for all assays included probe concentrations first tested at 100 nM, 200 nM, and 400 nM with an 800 nM primer concentration. Additional primer concentrations were tested at 400 nM, 1600nM, and 3200 nM. Annealing cycling conditions were tested at 55°C and 50°C for 30 and 20 cycles, respectively.
