## Supplemental Table 3 for "Simultaneous detection and quantification of multiple pathogen targets in wastewater"

**S3 Table.** qPCR Primer and Probe Sequences for TAC

| **Organism** | **Gene** | **Primer or probe sequence (5' - 3')** | **Reference** |
| --- | --- | --- | --- |
| astrovirus | Capsid | Fwd: CAGTTGCTTGCTGCGTTCA |  |
|  |  | Rev: CTTGCTAGCCATCACACTTCT | [1] |
|  |  | Probe: CACAGAAGAGCAACTCCATCGC |  |
| Pan-enterovirus | 5'UTR | Fwd: CCCTGAATGCGGCTAATCC |  |
|  |  | Rev: GCGATTGTCACCATWAGCAG | [1] |
|  |  | Probe: CCGACTACTTTGGGWGTCCGT |  |
| norovirus GI | ORF1-ORF2 | Fwd: CGYTGGATGCGNTTYCATGA |  |
|  |  | Rev: CTTAGACGCCATCATCATTYAC | [1] |
|  |  | Probe: TGGACAGGAGATCGC |  |
| norovirus GII | ORF1-ORF2 | Fwd: CARGARBCNATGTTYAGRTGGATGAG |  |
|  |  | Rev: TCGACGCCATCTTCATTCACA | [1] |
|  |  | Probe: TGGGAGGGCGATCGCAATCT |  |
| sapovirus (I, II, IV) | RdRp | Fwd: GAYCAGGCTCTCGCYACCTAC |  |
|  |  | Rev: CCCTCCATYTCAAACACTA | [1] |
|  |  | Probe: CYTGGTTCATAGGTGGTRCAG |  |
| sapovirus V | RdRp | Fwd: TTTGAACAAGCTGTGGCATGCTAC |  |
|  |  | Rev: CCCTCCATYTCAAACACTA | [1] |
|  |  | Probe: CAGCTGGTACATTGGTGGCAC |  |
| adenovirus 40/41 | Hexon b | Fwd: AACTTTCTCTCTTAATAGACGCC |  |
|  |  | Rev: AGGGGGCTAGAAAACAAAA | [1] |
|  |  | Probe: CTGACACGGGCACTCT |  |
| rotavirus | NSP3 | Fwd: ACCATCTWCACRTRACCCTCTATGAG |  |
|  |  | Rev: GGTCACATAACGCCCCTATAGC | [1] |
|  |  | Probe: AGTTAAAAGCTAACACTGTCAAA |  |
| *Campylobacter jejuni*/*C. coli* | *cadF* | Fwd: CTGCTAAACCATAGAAATAAAATTTCTCAC |  |
|  |  | Rev: CTTTGAAGGTAATTTAGATATGGATAATCG | [1] |
|  |  | Probe: CATTTTGACGATTTTTGGCTTGA |  |
| *Clostridioides difficile* | *tcdB* | Fwd: GGTATTACCTAATGCTCCAAATAG |  |
|  |  | Rev: TTTGTGCCATCATTTTCTAAGC | [1] |
|  |  | Probe: CCTGGTGTCCATCCTGTTTC |  |
| EAEC (aaiC) | *aaiC* | Fwd: ATTGTCCTCAGGCATTTCAC |  |
|  |  | Rev: ACGACACCCCTGATAAACAA | [1] |
|  |  | Probe: TAGTGCATACTCATCATTTAAG |  |
| EAEC (aatA) | *aatA* | Fwd: CTGGCGAAAGACTGTATCAT |  |
|  |  | Rev: TTTTGCTTCATAAGCCGATAGA | [1] |
|  |  | Probe: TGGTTCTCATCTATTACAGACAGC |  |
| STEC (stx1) | *stx1* | Fwd: ACTTCTCGACTGCAAAGACGTATG |  |
|  |  | Rev: ACAAATTATCCCCTGWGCCACTATC | [1] |
|  |  | Probe: CTCTGCAATAGGTACTCCA |  |
| STEC (stx2) | *stx2* | Fwd: CCACATCGGTGTCTGTTATTAACC |  |
|  |  | Rev: GGTCAAAACGCGCCTGATAG | [1] |
|  |  | Probe: TTGCTGTGGATATACGAGG |  |
| EPEC (eae) | *eae* | Fwd: CATTGATCAGGATTTTTCTGGTGATA |  |
|  |  | Rev: CTCATGCGGAAATAGCCGTTA | [1] |
|  |  | Probe: ATACTGGCGAGACTATTTCAA |  |
| EPEC (bfpA) | *bfpA* | Fwd: TGGTGCTTGCGCTTGCT |  |
|  |  | Rev: CGTTGCGCTCATTACTTCTG | [1] |
|  |  | Probe: CAGTCTGCGTCTGATTCCAA |  |
| ETEC LT | *LT* | Fwd: TTCCCACCGGATCACCAA |  |
|  |  | Rev: CAACCTTGTGGTGCATGATGA | [1] |
|  |  | Probe: CTTGGAGAGAAGAACCCT |  |
| ETEC ST | *STh* | Fwd: GCTAAACCAGYAGRGTCTTCAAAA  Rev: CCCGGTACARGCAGGATTACAACA  Probe: TGGTCCTGAAAGCATGAA | [1] |
|  | *STp* | Fwd: TGAATCACTTGACTCTTCAAAA |  |
|  |  | Rev: GGCAGGATTACAACAAAGTT |  |
|  |  | Probe: TGAACAACACATTTTACTGCT |  |
| EIEC/*Shigella* | *ipaH* | Fwd: CCTTTTCCGCGTTCCTTGA |  |
|  |  | Rev: CGGAATCCGGAGGTATTGC | [1] |
|  |  | Probe: CGCCTTTCCGATACCGTCTCTGCA |  |
| *Salmonella spp.* | *ttr* | Fwd: CTCACCAGGAGATTACAACATGG |  |
|  |  | Rev: AGCTCAGACCAAAAGTGACCATC | [1] |
|  |  | Probe: CACCGACGGCGAGACCGACTTT |  |
| *E. coli* O157: H7 | *rfbE* | Fwd: TTTCACACTTATTGGATGGTCTCAA |  |
|  |  | Rev: CGATGAGTTTATCTGCAAGGTGAT | [1] |
|  |  | Probe: CTCTCTTTCCTCTGCGGTCCT |  |
| *Cryptosporidium*  (pan-Crypto) | 18S rRNA | Fwd: GGGTTGTATTTATTAGATAAAGAACCA |  |
|  |  | Rev: AGGCCAATACCCTACCGTCT | [1] |
|  |  | Probe: TGACATATCATTCAAGTTTCTGAC |  |
| *Giardia* spp. | 18S rRNA | Fwd: GACGGCTCAGGACAACGGTT |  |
|  |  | Rev: TTGCCAGCGGTGTCCG | [1] |
|  |  | Probe: CCCGCGGCGGTCCCTGCTAG |  |
| *E. histolytica* | 18S rRNA | Fwd: ATTGTCGTGGCATCCTAACTCA |  |
|  |  | Rev: GCGGACGGCTCATTATAACA | [1] |
|  |  | Probe: TCATTGAATGAATTGGCCATTT |  |
| *Entamoeba* spp. | 18S rRNA | Fwd: AAACGATGTCAACCAAGGATTG |  |
|  |  | Rev: TCCCCCTGAAGTCCATAAACTC | [1] |
|  |  | Probe: CCTTGTTCAGAACTTAAAGAGAAA |  |
| *Ascaris lumbricoides* | *ITS1* | Fwd: GCCACATAGTAAATTGCACACAAAT |  |
|  |  | Rev: GCCTTTCTAACAAGCCCAACAT | [1] |
|  |  | Probe: TTGGCGGACAATTGCATGCGAT |  |
| *Trichuris trichiura* | 18S rRNA | Fwd: TTGAAACGACTTGCTCATCAACTT |  |
|  |  | Rev: CTGATTCTCCGTTAACCGTTGTC | [1] |
|  |  | Probe: CGATGGTACGCTACGTGCTTACCATGG |  |
| *Necator americanus* | ITS-2 | Fwd: CTGTTTGTCGAACGGTACTTGC |  |
|  |  | Rev: ATAACAGCGTGCACATGTTGC | [1] |
|  |  | Probe: CTGTACTACGCATTGTATAC |  |
| *Strongyloides stercoralis* | dispered repetitive sequence | Fwd: TCCAGAAAAGTCTTCACTCTCCAG |  |
|  |  | Rev: TGCGTTAGAATTTAGATATTATTGTTGCT | [1] |
|  |  | Probe: TCAGCTCCAGTTGAACAACAGCCTCCAA |  |
| *Ancylostoma duodenales* | ITS-2 | Fwd: GAATGACAGCAAACTCGTTGTTG |  |
|  |  | Rev: ATACTAGCCACTGCCGAAACGT | [1] |
|  |  | Probe: ATCGTTTACCGACTTTAG |  |
| *Enterobius vermicularis* | 5S rRNA | Fwd:  CAAACAACTGCATCACCAATAAC |  |
|  |  | Rev: AGTGTAGAGCAATAAGCAGTAAAG | [2] |
|  |  | Probe:  TACCAACAACACTTGCACGTCTCTTCA |  |
| *Hymenolepis nana* | ITS1 | Fwd: CATTGTGTACCAAATTGATGATGAGTA |  |
|  |  | Rev: CAACTGACAGCATGTTTCGATATG | [3] |
|  |  | Probe: CGTGTGCGCCTCTGGCTTACCG |  |
| 16s  **Used in Liu et al. 2013 |  | Fwd: TGCAAGTCGAACGAAGCACTTTA |  |
|  |  | Rev: GCAGGTTACCCACGCGTTAC | [1] |
|  |  | Probe: CGCCACTCAGTCACAAA |  |
| Phocine herpesvirus (PhHV) | gB | Fwd: GGGCGAATCACAGATTGAATC |  |
|  |  | Rev: GCGGTTCCAAACGTACCAA | [1] |
|  |  | Probe: TATGTGTCCGCCACCATCT |  |
| *Yersinia enterocolitica* | *lytA* | Fwd:  TGATTCACCAGCAGCAATAC |  |
|  |  | Rev: GGCATCATGAAAGGCGG | [1] |
|  |  | Probe: TGTCGGTTTCTCCTTCCAGG |  |
| *Heliobacter pylori* | *ureC* | Fwd: GACACCAGAAAAAGCGGCTA |  |
|  |  | Rev:  AGCGCATGTCTTCGGTTAAA | [1] |
|  |  | Probe: TCACTAAAGCGTTTTCTACC |  |
| *Plesiomonas shigelloides* | *gyrB* | Fwd: CCGCCGTGAAGGCAAAG |  |
|  |  | Rev: GCTACCGGCTCACCCAGAT | [1] |
|  |  | Probe: CACACCCAAGAATAC |  |
| *Cyclospora cayetanensi* | 18s rRNA | Fwd:  AAAAGCTCGTAGTTGGATTTCTG |  |
|  |  | Rev: AACACCAACGCACGCAGC | [1] |
|  |  | Probe: AAGGCCGGATGACCACGA |  |
| *Cystoisospora belli* | 18S rRNA | Fwd: ATATTCCCTGCAGCATGTCTGTTT |  |
|  |  | Rev: CCACACGCGTATTCCAGAGA | [1] |
|  |  | Probe: CAAGTTCTGCTCACGCGCTTCTGG |  |
| *Blastocystis* spp. | 18S rRNA | Fwd: TGGTCCGRTGAACACTTTGGAT |  |
|  |  | Rev: CCTACGGAAACCTTGTTACGACTTCA | [1] |
|  |  | Probe: CTTCCTCTAAATGRTAAGATT |  |
| *Enterocytozoon bieneusi* | SSU rRNA | Fwd: TGTGTAGGCGTGAGAGTGTATCTG |  |
|  |  | Rev: CATCCAACCATCACGTACCAATC | [1] |
|  |  | Probe: CACTGCACCCACATCCCTCACCCTT |  |
| *Encephalitozoon intestinalis* | ITS | Fwd: CACCAGGTTGATTCTGCCTGAC |  |
|  |  | Rev: CTAGTTAGGCCATTACCCTAACTACCA | [1] |
|  |  | Probe: CTATCACTGAGCCGTCC |  |
| *Balantidium coli* | ITS-1 | Fwd: TGCAATGTGAATTGCAGAACC |  |
|  |  | Rev: TGGTTACGCACACTGAAACAA | [4] |
|  |  | Probe: CTGGTTTAGCCAGTGCCAGTTGC |  |
| *Acanthamoeba* spp. | 18S rRNA | Fwd: CCCAGATCGTTTACCGTGAA |  |
|  |  | Rev: TAAATATTAATGCCCCCAACTATC | [5] |
|  |  | Probe: CTGCCACCGAATACATTAGCATGG |  |
| hepatitis A | 5' NCR | Fwd: TCACCGCCGTTTGCCTAG |  |
|  |  | Rev: GGAGAGCCCTGGAAGAAAG | [6] |
|  |  | Probe: TTAATTCCTGCAGGTTCAGG |  |
| MS2 | *MS2g1* | Fwd: TGGCACTACCCCTCTCCGTATTCAC  Rev: GTACGGGCGACCCCACGATGAC  Probe: CACATCGATAGATCAAGGTGCCTACAAGC | [1] |
| SARS-CoV-2 | N1 gene | Fwd: GACCCCAAAATCAGCGAAAT |  |
|  |  | Rev: TCTGGTTACTGCCAGTTGAATCTG | [7] |
|  |  | Probe: ACCCCGCATTACGTTTGGTGGACC |  |
