## Supplemental Table 2 for "Simultaneous detection and quantification of multiple pathogen targets in wastewater"

**S2 Table. Matched samples comparison on TAC for pathogen types**

|  |  | ****Concentrating Pipette Pellet**** | | ****Direct Extraction**** | | ****Skim Milk Pellet**** | |
| --- | --- | --- | --- | --- | --- | --- | --- |
| **Pathogen Type** | **N** | ***No Detection*** | ***Detection*** | ***No Detection*** | ***Detection*** | ***No Detection*** | ***Detection*** |
| **Bacteria** | **17** | **7 (41%)** | **10 (59%)** | **15 (88%)** | **2 (12%)** | **6 (35%)** | **11 (65%)** |
| **Fungus/Algae** | **3** | **0** | **3 (100%)** | **2 (67%)** | **1 (33%)** | **1 (33%)** | **2 (67%)** |
| **Helminth** | **7** | **7 (100%)** | **0** | **7 (100%)** | **0** | **7 (100%)** | **0** |
| **Protozoa** | **8** | **4 (50%)** | **4 (50%)** | **5 (63%)** | **3 (38%)** | **5 (63%)** | **3 (38%)** |
| **Virus** | **7** | **2 (29%)** | **5 (71%)** | **4 (57%)** | **3 (43%)** | **0** | **7 (100%)** |

**SMP methods detected viral target amplification 100% (n=7/7), where CPP methods detected 71% (5/7), and direct extraction 43% (3/7).**

**Of the four matched samples, direct extraction only resulted in detection for 25% of any single TAC target, whereas SMP and CPP performed at 52% and 54%, respectively [data not shown].**
